# Unveiling the Burden of MASLD and Liver Fibrosis in India: Novel Insights from the Phenome India Study into fibrosis without MASLD

**DOI:** 10.1101/2025.10.11.25337761

**Authors:** Meghana Arvind, Anshul Verma, K Sreeshma Raj, Satyartha Prakash, Vignesh S Kumar, Mohammad Azhar Uddin, Ayushi Narayan, Mamta Rathore, Nancy Rawat, Ankita Sahu, Yogesh Kumar, Pulkit Hasmukhbhai Leuva, Monika Sharma, S Rajesh, Dwaipayan Saha, Ankita Mridha, Ishant Jyoti Nath, Ashique Hussain, Borsha Rajkumari, Mamta Thapa, Neha Kumari, S Vishwapriya, Shilpak Chatterjee, Dipyaman Ganguly, Ashish Awasthi, Vamsi Yenamandra, Ajay Pratap Singh, Aastha Mishra, Swasti Raychaudhuri, Karthik Bharadwaj Tallapaka, Giriraj Ratan Chandak, Mahesh J Kulkarni, Mahesh Dharne, Romi Wahengbam, Umakanta Subudhi, Sagnik Biswas, Shalimar, Phenome India Consortium, Kumardeep Chaudhary, Shantanu Sengupta, Partha Chakraborty, Viren Sardana

## Abstract

**Background:** Metabolically–dysfunction–associated steatotic liver disease (MASLD) is rising globally, including in India, yet community-based data remain scarce. We addressed this critical knowledge gap by assessing the prevalence, distribution, and characteristics of MASLD subgroups and fibrosis, leveraging the Phenome India–CSIR Health Cohort Knowledgebase (PI-CHeCK) study.

**Methods:** In this prospective, nationwide study, we recruited 10267 adults across 37 laboratories of the Council of Industrial Research (CSIR) from 27 Indian cities. Steatosis and fibrosis were assessed by Transient Elastography using Fibroscan, with associated clinical, biochemical, cytokine, anthropometric, and lifestyle data collected. Overall, crude and age-adjusted prevalence rates were estimated in the study population and various subgroups.

**Findings:** Of the 10267 individuals screened, 7764 were eligible for analysis after exclusions. Among these, 3,712 (47.8%) fulfilled MASLD criteria, corresponding to an age-adjusted prevalence of 36.3%. Significant fibrosis (≥F2) was more frequent in MASLD (6.3% [234 of 3688]) than in cases of cryptogenic fibrosis without MASLD (1.7% [69 of 4027]), corresponding to an age-adjusted prevalence of 4.1% in MASLD. Overall age-adjusted prevalence of significant fibrosis was 2.3% in the entire cohort, which clustered in older adults (>60 years) and in those with diabetes or obesity II, with evidence of regional variation, peaking in Jorhat, Assam. Importantly, fibrosis in participants without MASLD emerged as a distinct subgroup with disproportionately elevated cytokine levels, exceeding those in MASLD with fibrosis, suggesting a cytokine-rich, high-risk phenotype.

**Conclusion:** In this nationwide Indian cohort, MASLD affected over one-third of participants with a substantial burden of fibrosis. Notably, fibrosis without-MASLD emerged as a cytokine-rich, high-risk phenotype, underscoring an underrecognized dimension of liver disease with major public health implications.

## Introduction

Metabolic-dysfunction-associated steatotic liver disease (MASLD), previously known as non-alcoholic fatty liver disease (NAFLD), characterised by excessive intrahepatic lipid accumulation, is a major global health concern, given its association with substantial morbidity and mortality (1). Globally, MASLD affects approximately 25–50% of the adult population, with prevalence rates influenced by ethnicity, dietary patterns, socioeconomic status and genetic factors (2, 3). Recent projections also suggest that the prevalence of obesity is expected to rise among men and women from 342 million (337–346) and 496 million (490–503) in 2021 to 838 million (692–921) and 1.11 billion (0.942–1.21) in men and women, respectively, by the year 2050 (4). In India, while there are no major studies available for community-wide prevalence of MASLD in the general population, the prevalence of NAFLD is alarmingly high, ranging from 9–32% in urban areas, with certain regions reporting a prevalence of nearly 60%, attributed to rapid urbanisation, sedentary lifestyles, and dietary shifts toward processed, calorie-dense foods (2, 5, 6). The South Asian population’s propensity for central obesity and genetic factors, such as polymorphisms in the PNPLA3 gene, further increases the susceptibility to obesity and, consequently, NAFLD (7, 8). NAFLD may progress to metabolic dysfunction-associated steatohepatitis (NASH), cirrhosis, and hepatocellular carcinoma(8). Fibrosis is a strong predictor of adverse outcomes in MASLD, determining risk of cirrhosis, hepatocellular carcinoma, liver-related mortality, and extrahepatic complications (9). Recent studies indicate that 10–15% of individuals with NAFLD or MASLD may already have advanced fibrosis, yet most remain undiagnosed until decompensation (7, 8, 10–12). Importantly, significant fibrosis is also observed in individuals without hepatic steatosis, indicating that liver fat alone may be an insufficient screening marker (13). Thus, population-level screening for fibrosis—irrespective of fatty liver status—may therefore be essential for risk stratification and timely intervention.

The asymptomatic nature of early-stage MASLD often leads to a delayed diagnosis, with patients being detected only at advanced stages of liver disease. This not only limits the therapeutic measures, but is also associated with higher non-liver related mortality (cardiac disease, extrahepatic malignancies) (14). The delay reduces opportunities for preventive interventions in individuals at risk of developing MASLD. An integral component of developing and implementing such preventive measures is the availability of nationwide data that provide reliable estimates of MASLD, prevalence of cardiometabolic risk factors (CMRF), and stage of disease at the community level. However, most studies on the prevalence of MASLD in India are hospital-based, drawn from endocrinology clinics, and gastroenterology or hepatology units at tertiary care centres, and may be subject to referral bias and suffer from the “iceberg” effect (15, 16). Heterogeneity in study populations, diagnostic criteria, and methodologies further complicates comparisons across reports.

The Phenome India cohort offers a unique opportunity to address these gaps. This nationwide, community-based urban and semi-urban cohort of working and retired employees and their spouses is relatively homogeneous in terms of socioeconomic and educational background, yet is geographically diverse across 27 cities, enabling robust, generalizable estimates of MASLD and fibrosis throughout India(17).

Thus, in this study, we report the prevalence and distribution of MASLD and fibrosis in more than 7,700 participants from the Phenome India cohort. Notably, beyond the expected high burden of MASLD fibrosis, we identify a distinct subgroup of without-MASLD fibrosis with disproportionately elevated cytokine levels—a cytokine-rich, high-risk phenotype representing an underrecognized dimension of liver disease in India.

## Participants and Methods

### Study design

We used the Phenome India–CSIR Health Cohort Knowledgebase (PI-CHeCK), a multi-centre longitudinal cohort established by the Council of Scientific and Industrial Research (CSIR), to estimate MASLD prevalence, grade community-level disease severity, and profile associated cardiometabolic risk factors (CMRF/s) (18) (17). PI-CHeCK includes 37 CSIR laboratories and affiliated centres across 17 states and 2 Union Territories in India. The study was approved by the Institutional Human Ethics Committee (IHEC) at CSIR-IGIB [Institute of Genomics and Integrative Biology] (reference number: CSIR-IGIB/IHEC/2023-24/16) and registered with the Clinical Trials Registry of India (CTRI/2024/01/061807). The study complies with the Declaration of Helsinki, and written informed consent was obtained from the participants. The data collection was carried out between December 2023 to June 2024.

### Inclusion and Exclusion criteria

All permanent staff members of CSIR, including current working employees, retirees, and their spouses, who responded to the recruitment campaign and provided voluntary consent, were considered for participation in the Phenome India Cohort. Exclusion criteria were age <18 years, pregnancy and unwillingness to participate.

Specific exclusion criteria applied to the analysis reported in this work were age <18 years, any alcohol use, pregnancy and unwillingness to participate. Hepatitis B and Hepatitis C were tested for those who had CAP≥248 and if CAP was <248, then with LSM≥6.5 and positives were excluded from analysis.

### Transient Elastography

After overnight fasting, hepatic steatosis and fibrosis were measured using FibroScan FS Mini 430 Plus^TM^ (Echosens, Paris, France) with M or XL probes based on body mass index (BMI), guided by the auto-recommendation system of the machine (19, 20). Liver Stiffness Measurement (LSM) and Continuous Attenuation Parameter (CAP) measurements were performed by trained personnel blinded to participants’ clinical data and study outcomes. For each participant, we obtained ten consecutive valid readings for LSM with an Interquartile range/median (IQR/M) ratio <0.3. Failure of transient elastography was defined as the inability to obtain a valid LSM or CAP measurement (19, 20). Contraindications for doing Fibroscan have been mentioned in previously published work (17).

### MASLD

MASLD was defined as hepatic steatosis detected on transient elastography (CAP value >248 dB/m,(5, 21)) with any one or more of five CMRFs: obesity, dysglycemia, hypertension (HTN) or HDL levels or triglyceride levels (21, 22).

### Hepatic Steatosis

Hepatic steatosis was defined as CAP value >248 dB/m, with severity graded as: S0 (no steatosis)-CAP <249 dB/m; S1 (mild steatosis)-CAP 249–267 dB/m; S2 (moderate steatosis)-CAP 268–279 dB/m; S3 (severe steatosis)-CAP ≥280 dB/m (5).

### Hepatic Fibrosis

LSM assessed on TE was correlated with hepatic fibrosis according to the METAVIR scoring system: F0/F1 (no fibrosis/mild fibrosis)-LSM <8.2 kPa; F2 (moderate fibrosis)-LSM 8.2 to <9.7 kPa; F3 (severe fibrosis)-LSM 9.7 to <13.6 kPa; F4 (cirrhosis)-LSM ≥13.6 kPa (23).

### Definitions of CMRFs for assessing MASLD status

#### 1. Dysglycaemia

Dysglycaemia was labelled based on one or more of the following criteria: fasting blood glucose (FBG) ≥100 mg/dL, glycated haemoglobin (HbA1c) ≥5.7%, a self-reported history of Type 2 diabetes, or self-reported use of medication for Type 2 diabetes control (5, 21, 22).

#### 2. Overweight and Obesity

Body mass index ≥ 23 kg/m2 (Asian ethnicity) and Waist circumference ≥ 90 cm in men and ≥ 80 cm in women (South Asians and Chinese) (21).

#### 3. Blood Pressure (BP) Measurement

Systolic (SBP) and diastolic blood pressure (DBP) were measured using an automated electronic sphygmomanometer. Hypertension (HTN) was defined as SBP ≥130 or DBP ≥85 mmHg, or a self-reported diagnosis or use of antihypertensive drugs(21).

#### 4. Triglyceride Levels

Plasma triglycerides ≥150 mg/dl or lipid-lowering treatment (21).

#### 5. HDL Levels

HDL-cholesterol ≤ 39 mg/dl in men and ≤ 50 mg/dl in women, or lipid-lowering treatment (21).

## General Definitions

### Overweight and Obesity

BMI was used to categorize weight status according to the Asia-Pacific threshold values as follows: underweight: BMI <18.5 kg/m²; normal/healthy weight: BMI ≥18.5 and <23 kg/m²; overweight: BMI ≥23 and <25 kg/m²; obesity class I: BMI ≥25 and <30 kg/m²; obesity class II: BMI ≥30.0 kg/m² (5, 21, 24).

### Type 2 Diabetes Mellitus (T2DM)

Type 2 diabetes mellitus (T2DM) was labelled based on one or more of the following criteria: fasting blood glucose (FBG) ≥126 mg/dL, glycated haemoglobin (HbA1c) ≥6.5%, a self-reported history of Type 2 diabetes, or self-reported use of medication for Type 2 diabetes control (5).

### Prediabetes

Prediabetes was labelled based on one or more of the following criteria: fasting blood glucose (FBG) ≥100 and ≤ 125 mg/dL, glycated haemoglobin (HbA1c) ≥5.7 and ≤ 6.4%.

### Elevated Blood Pressure

Elevated Blood Pressure was defined as SBP ≥130 or DBP ≥85 mmHg, or a self-reported diagnosis or use of antihypertensive drugs (21).

### Dyslipidemia

Total Cholesterol ≥ 200 mg/dl or Serum triglycerides levels ≥ 150 mg/dl (5) or lipid-lowering treatment.

### Cytokine Analysis

A Bio-Rad Human Cytokine 48-Plex assay fibrosis (Cat. No. 12007283, Bio-Rad Laboratories, USA) was used for cytokine profiling, stratifying patients into four clinical subgroups: MASLD with/without fibrosis, and without-MASLD with/without fibrosis, following the manufacturer’s recommended protocol (Supplementary Methods). The assay was performed at two Institutes; hence, data were normalized (Supplementary methods) (25).

### Anthropometry

Anthropometry was carried out to measure height, weight, chest (CC), waist (WC), abdominal (AC), and hip circumferences (HC). Height (in cm) (Supplementary Methods) (26, 27). Details of anthropometry, biochemistry, and other tests done are available in the protocol paper of Phenome India, published previously (17).

### Statistical analysis

Normality of data was assessed using Shapiro-Wilk’s test and Q-Q plots. Comparisons of continuous variables between two or three independent groups were performed using the Mann-Whitney U test or Kruskal-Wallis, respectively. Categorical variables were compared using the Chi-square test or Fisher’s exact as appropriate. Cytokine-based statistical analysis was performed for non-normally distributed data. Data has been represented as Median (IQR range) or n (%) mostly unless specified otherwise.

A Python Program was utilised with the required libraries for large-scale data analysis and statistical computation for adjusted value. The script used the Benjamini-Hochberg (BH) procedure for False Discovery Rate (FDR) correction to adjust p-values, to control the FDR, which is the expected proportion of false positives among all significant results. Iterative p-values were computed when the sample size difference between the two groups was greater than 4-fold. Graph Pad Prism and Python were utilised for data visualisation. Multivariate Regression Analysis was done using Stata 18.5 and 19.

Age-adjusted values were calculated against the urban population data of India from the Census 2011 (28).

## Results

### Demographics

The Phenome India cohort included 10267 participants recruited from 27 cities across India between December 2023 and June 2024. Of these 7764, were eligible for analysis (Figure 1). Of these 7764, 47.3% were males; median age 52 (IQR 41–62) years and 52.7% were females; median age 49 (IQR 40 –59) years. Among 7762 participants with recorded height and weight, 18% had a normal or low BMI (lean), while 82% of participants were categorized as overweight or obese (BMI ≥ 23 kg/m²) (Table 1). Among males, 22.2% participants were lean, in contrast to 14.43% females who were categorized as lean. A greater number of males were either overweight or had Grade 1 obesity, while the prevalence of Grade 2 obesity was higher in females. Overall demographics for all 10267 participants are given in the Supplementary Table 1.

**Figure 1:**
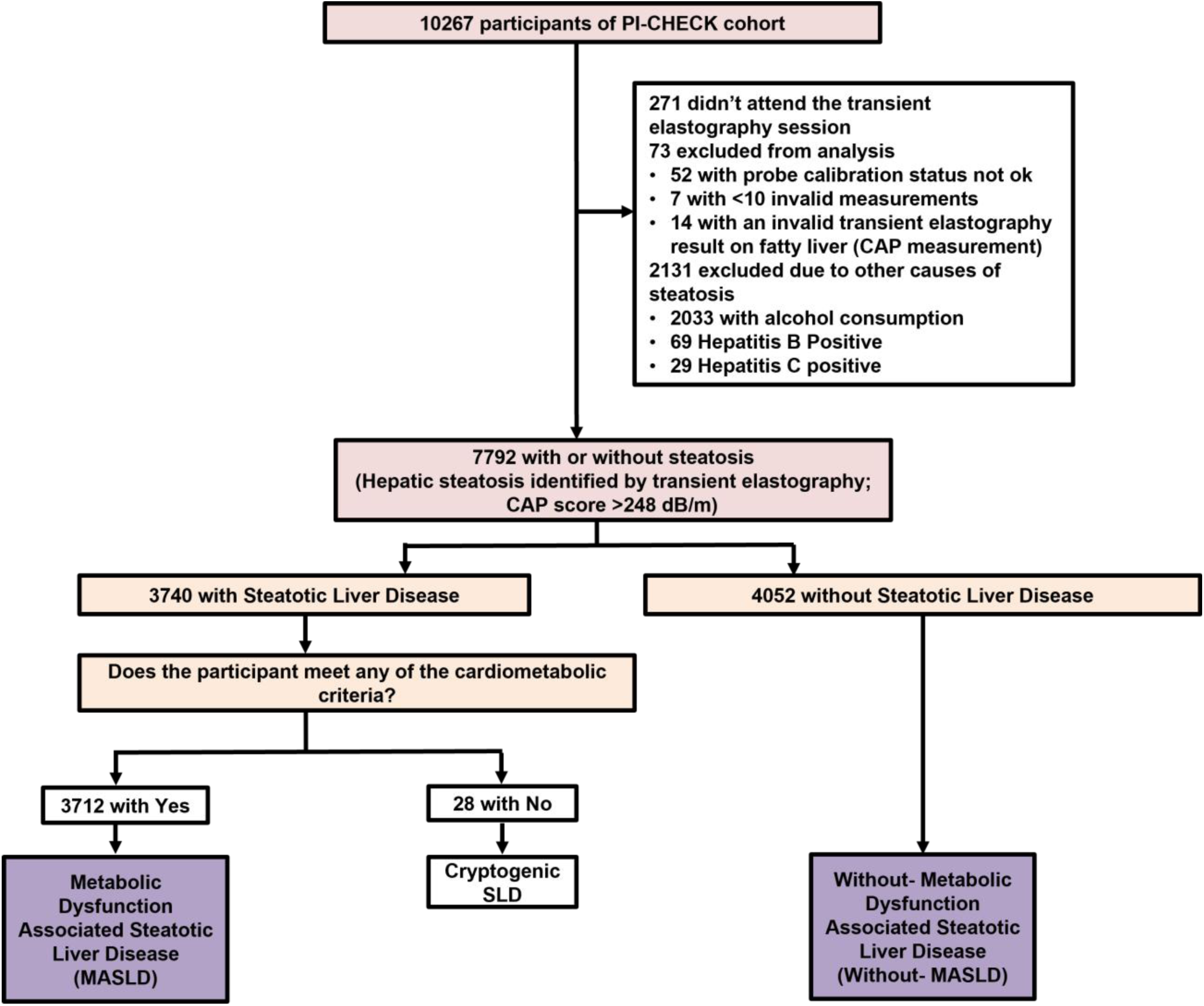
Flow for identification of MASLD participants. Abbreviations: CAP, Controlled Attenuation Parameter; SLD, Steatotic Liver Disease.

**Table 1:** Demographics of samples analysed for MASLD and without MASLD (n=7764)

| Variable | Total | Male | Female |
| --- | --- | --- | --- |
| Sample size | 7764 (100.0%) | 3671 (47.3%) | 4093 (52.7%) |
| **Demographics and Biochemistry** |  |  |  |
| Age (years) | 50.00 (41.00 - 61.00) | 52.00 (41.00 - 62.00) | 49.00 (40.00 - 59.00) |
| AST (u/L) | 24.10 (20.20 - 29.70) | 25.90 (21.90 - 31.80) | 22.70 (19.10 - 27.60) |
| ALT (u/L) | 21.90 (16.10 - 31.40) | 26.00 (19.20 - 37.30) | 18.90 (14.40 - 26.10) |
| Glucose Fasting (mg/dL) | 90.80 (84.30 - 102.20) | 91.90 (85.20 - 104.30) | 89.60 (83.50 - 100.50) |
| Glycosylated Hemoglobin (%) | 5.60 (5.30 - 6.20) | 5.70 (5.30 - 6.30) | 5.60 (5.30 - 6.10) |
| Diabetes | 1927 (24.8%) | 1005 (27.4%) | 922 (22.5%) |
| Homa IR | 1.87 (1.27 - 2.74) | 1.92 (1.29 - 2.84) | 1.82 (1.25 - 2.65) |
| Total cholesterol (mg/dL) | 176.00 (150.90 - 202.40) | 174.20 (149.10 - 200.55) | 177.80 (153.20 - 204.50) |
| Serum HDL cholesterol (mg/dL) | 48.10 (42.00 - 55.20) | 45.10 (39.70 - 51.00) | 51.20 (44.90 - 58.80) |
| Serum LDL cholesterol (mg/dL) | 114.90 (95.30 - 135.43) | 115.20 (95.10 - 136.20) | 114.60 (95.40 - 134.90) |
| Serum Triglycerides (mg/dL) | 112.60 (84.47 - 151.22) | 119.70 (90.70 - 161.85) | 106.50 (80.00 - 141.30) |
| Hb (gm) | 13.10 (12.00 - 14.30) | 14.30 (13.40 - 15.00) | 12.20 (11.40 - 13.00) |
| Elevated BP | 4272 (55.0%) | 2160 (58.8%) | 2112 (51.6%) |
| **Anthropometry** |  |  |  |
| Weight (cm) | 67.30 (60.20 - 75.10) | 70.60 (63.80 - 78.60) | 64.10 (57.80 - 71.70) |
| Height (cm) | 160.00 (153.50 - 167.00) | 167.20 (162.88 - 171.70) | 154.10 (150.10 - 158.40) |
| Waist circumference | 87.53 (81.00 - 94.67) | 88.67 (83.43 - 95.17) | 85.67 (78.53 - 94.07) |
| WHR | 0.89 (0.83 - 0.95) | 0.93 (0.89 - 0.97) | 0.85 (0.80 - 0.90) |
| **BMI Categories** |  |  |  |
| Lean | 1405 (18.1%) | 814 (22.2%) | 591 (14.4%) |
| Overweight | 1446 (18.6%) | 852 (23.2%) | 594 (14.5%) |
| Obese I | 3487 (44.9%) | 1622 (44.2%) | 1865 (45.6%) |
| Obese II | 1424 (18.3%) | 383 (10.4%) | 1041 (25.4%) |

Continuous variables are summarized using median (Q1, Q3), and categorical variables are summarized using n (%); missing data for each variable were excluded when producing these summaries. Lean includes underweight: BMI <18.5 kg/m<sup>2</sup> and normal/healthy weight: BMI 18.5–22.9 kg/m<sup>2</sup>.
Abbreviations: Homa IR, Homeostatic Model Assessment of Insulin Resistance; HDL, High-density lipoprotein; LDL, Low-density lipoprotein; CAP, Controlled Attenuation Parameter.

Various anthropometric and biochemical parameters of the participants in the overall cohort, along with gender wise distribution, are shown in Table 1.

### Prevalence of MASLD

Of the 9923 individuals with valid TE measurements. 7764 participants remained after exclusion of participants who gave a history of any alcohol consumption and had positive coexistent liver disease (viral markers positive), of these, 3740 had SLD (CAP value >248). 3712 of 3740 met the diagnostic criteria for MASLD (Figure 1), with an overall prevalence of 47.8% (3712 of 7764, 95% CI: 46.6-48.9) (Figure 1) and an age-adjusted prevalence of 36.3%. We observed 45.9% of females and 49.9% of males as a crude percentage and 30.8% and 43.3% age-adjusted prevalence, respectively, in our pan-India cohort to be affected by MASLD. Crude age-wise and gender prevalence are provided in Supplementary Figure 1.

The prevalence of elevated BP (64.3% vs 46.5%, p<0.01) and Type 2 diabetes mellitus (T2DM) (34.5% vs 16.0%, p<0.01) was found to be markedly higher in individuals with MASLD than in those without MASLD. Individuals with MASLD also had higher median HbA1c values: 5.9 (5.5-6.6) vs 5.5 (5.2-5.8) [p<0.01], greater insulin resistance (IR) as measured by homeostatic model for insulin resistance (HOMA-IR): 2.3 (1.6-3.4) vs 1.4 (1.0-2.1) [p<0.01] and higher prevalence of dyslipidemia (57.4% vs 41.3%, p<0.01) compared to those with MASLD (Table 2). More expectedly, a significant difference was noted in the anthropometric parameters of individuals with MASLD as compared to those without MASLD (Table 3).

**Table 2:**
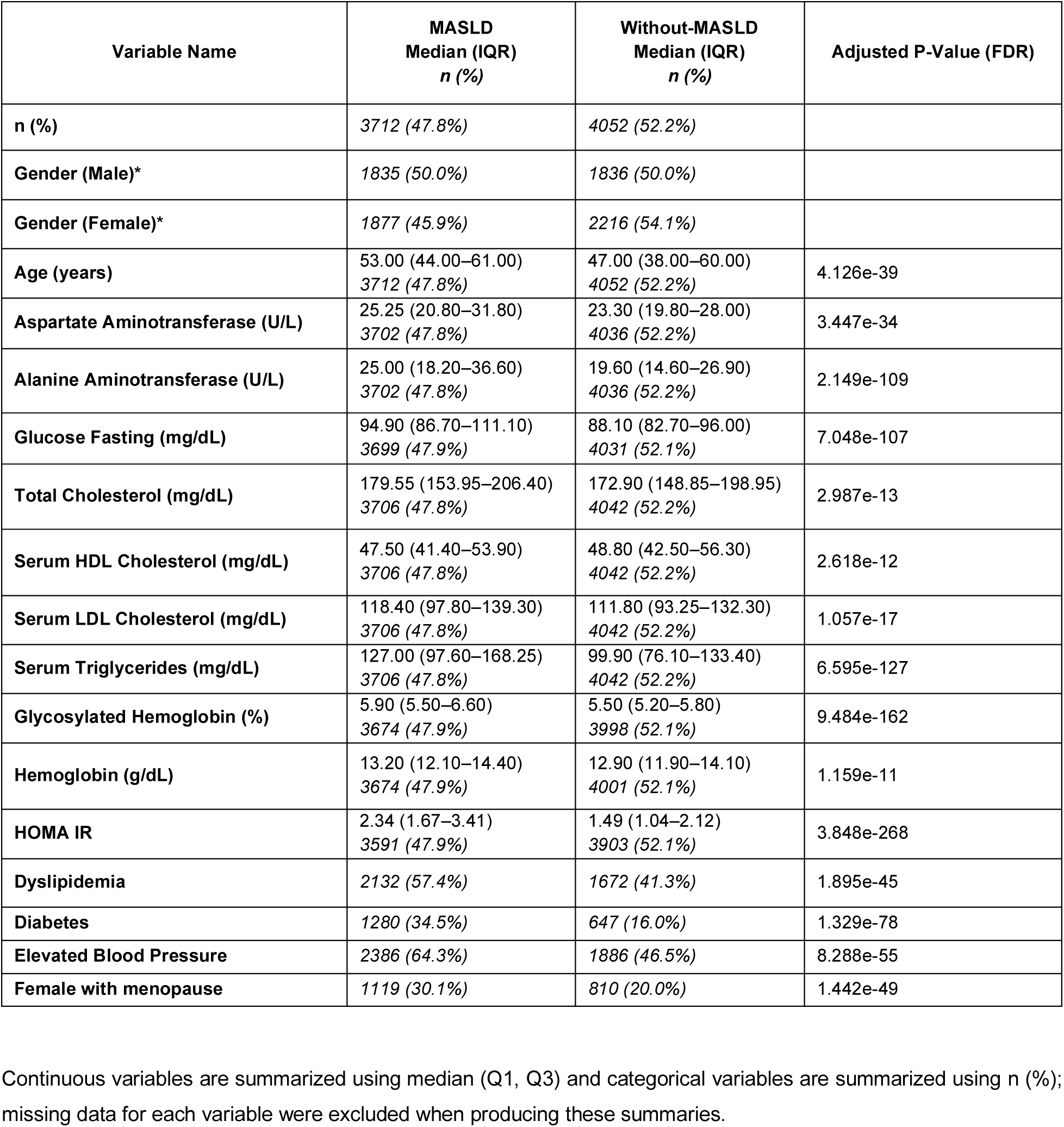
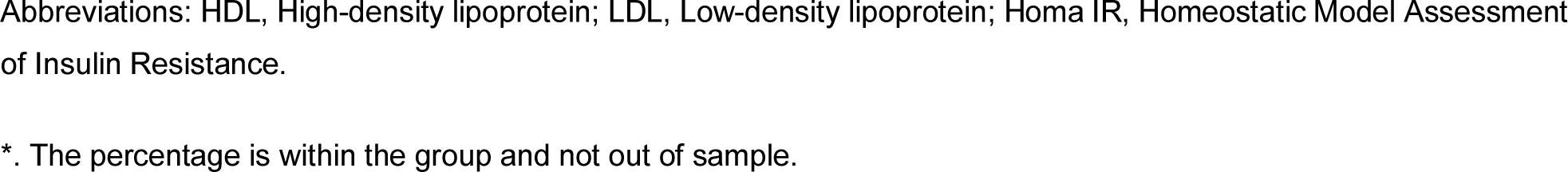
Comparison of Biochemical and clinical parameters between MASLD and without-MASLD population subgroups.

**Table 3:**
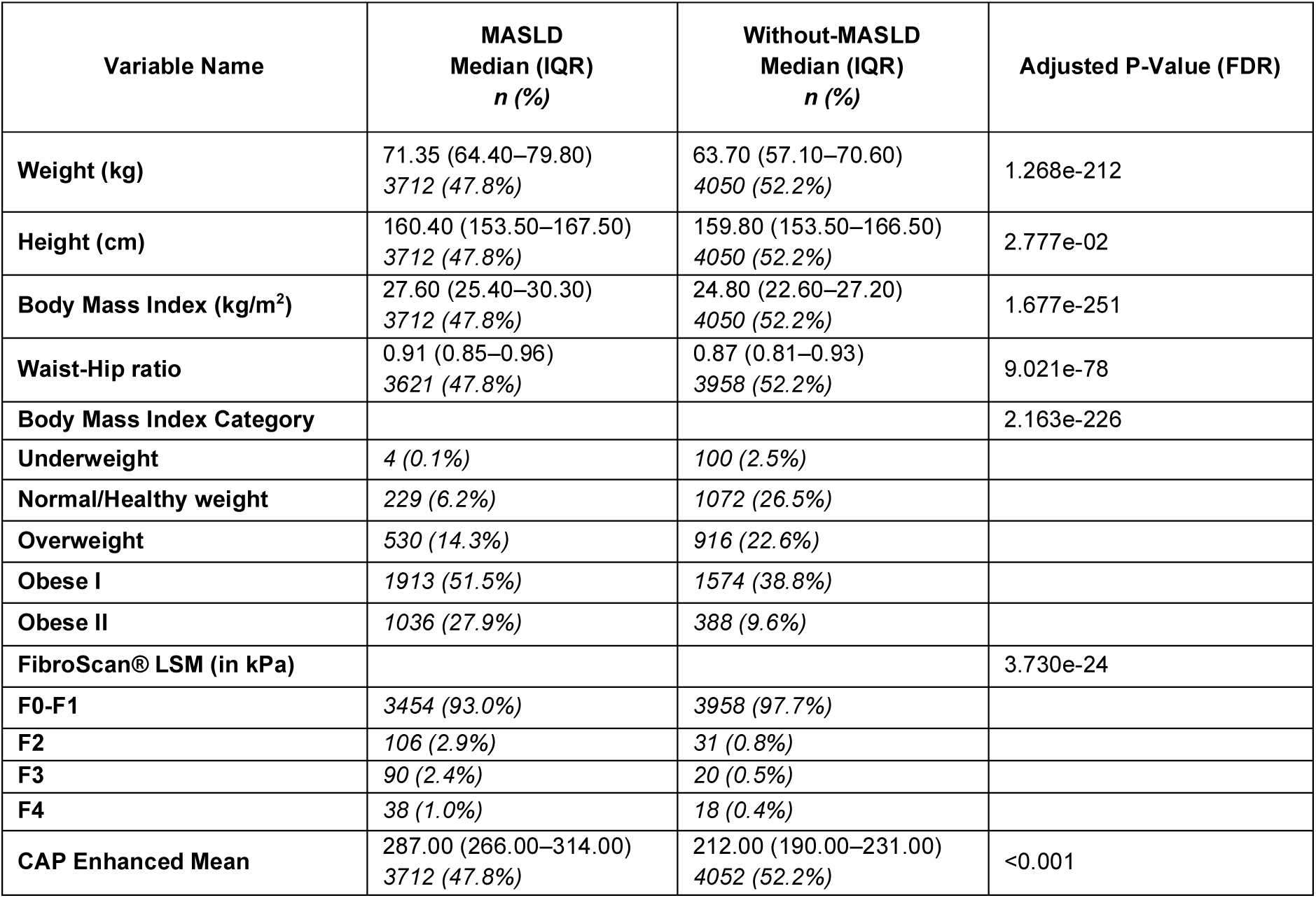
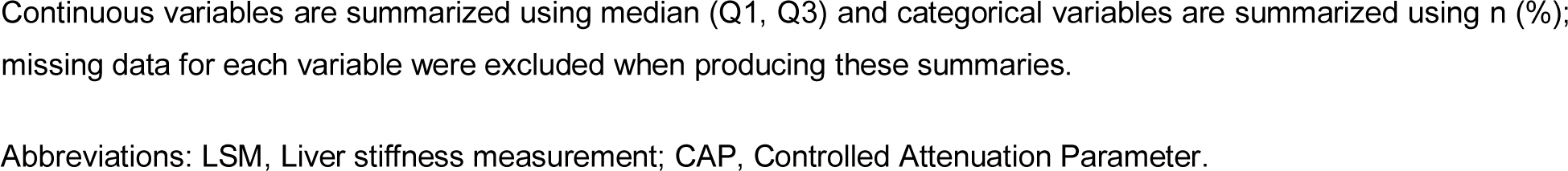
Comparison of anthropometric parameters and TE parameters between MASLD and the without-MASLD population subgroups.

There was no significant difference in the monthly income, dietary habits and educational status among individuals based on the presence or absence of MASLD (Supplementary Table 2). However, the number of females with menopause was higher in the MASLD group than in the group without MASLD (Table 2).

The MASLD group had significantly higher BMI (overweight and obese) individuals (93.7% vs 71%). The crude prevalence of significant liver fibrosis (≥F2) was markedly higher in the MASLD group (6.3%) as compared to the without-MASLD group (1.7%) (Table 3).

### Regression modelling for MASLD

Multivariate logistic regression identified obesity as the strongest risk factor for MASLD in India, with odds ratios (OR) rising steeply from overweight (OR 2.6, 95% CI 2.2–3.1) to Obese I (OR 5.6, 95% CI 4.6–6.8) and Obese II (OR 13.7, 95% CI 11.4–16.5). Central adiposity (waist–hip ratio ≥0.80 in females and 0.90 in males) was also independently associated (OR 1.6, 95% CI 1.4–1.8). Diabetes (OR 2.2, 95% CI 1.9–2.5), dyslipidemia (OR 1.6, 95% CI 1.5–1.8) and hypertension (OR 1.2, 95% CI 1.1–1.4) further increased MASLD risk (Supplementary table 3). Age showed a graded association, peaking at 50– 60 years (OR 1.7, 95% CI 1.3–2.3), while female gender was protective (OR 0.60, 95% CI 0.52–0.7). Notably, underweight status conferred reduced risk (OR 0.2, 0.06–0.6). Obesity is thus the predominant driver of MASLD in India, with additional contributions from diabetes, dyslipidemia, hypertension, and central adiposity, underscoring the need for aggressive metabolic risk management (Supplementary Table 3).

### Prevalence of MASLD across BMI

Among 3,712 MASLD participants stratified by BMI (lean 6.3%, overweight 14.3%, obese 79.4%), marked demographic and biochemical differences were observed (Supplementary Table 4). Lean MASLD individuals were older (median 58 years) compared to overweight (53 years) and obese (52 years; p<0.01). Sex distribution amongst these three groups differed substantially. Women comprise 55.3% of obese MASLD, with males being less at 44.7%. However, in the lean and overweight group, females were less at 36.1% and 30.8% respectively, and males predominated. Insulin resistance increased progressively with BMI: median HOMA-IR rose from 1.8 in lean to 2.5 in obese (p<0.01). HbA1c values varied minimally (6.0% vs. 5.8% vs. 5.9%), while fasting glucose was paradoxically highest in lean MASLD (99 mg/dL vs. 94 mg/dL in overweight/obese; p<0.01).

Lipid parameters were broadly comparable across groups; HDL was modestly lower in overweight/obese than lean (p<0.01), whereas triglycerides were slightly higher in lean without significance. Lean MASLD had the greatest burden of diabetes (42.5%) and elevated BP (67.0%), despite lower adiposity. In contrast, obesity was associated with a higher prevalence of post-menopausal women (32.2%). Together, these data suggest that lean MASLD represents a clinically distinct phenotype, characterized by older age, greater cardiometabolic burden, and adverse glycemic traits despite lower BMI.

### Geographical Distribution of MASLD in this cohort

The age-adjusted prevalence of MASLD demonstrated wide regional variation, ranging from ∼25% in Thiruvananthapuram and ∼30% in Kolkata, Dehradun and Durgapur to nearly 50% in Roorkee, Bhopal and Bhavnagar (Figure 2 and Supplementary Table 5). Major metropolitan centers such as Delhi, Bengaluru, Pune, Hyderabad and Chennai showed intermediate prevalence (∼35–45%). Men were consistently more affected than women (43.3% vs. 30.8%), with male prevalence exceeding 50% in many of the places. While extreme values were noted in Ghaziabad and Srinagar, these were based on small sample sizes and should be interpreted cautiously. Overall, the data reveal low prevalence in southern and eastern cities and higher burdens in several central and western regions, underscoring significant geographic and sex-specific heterogeneity in MASLD distribution across India.

**Figure 2:**
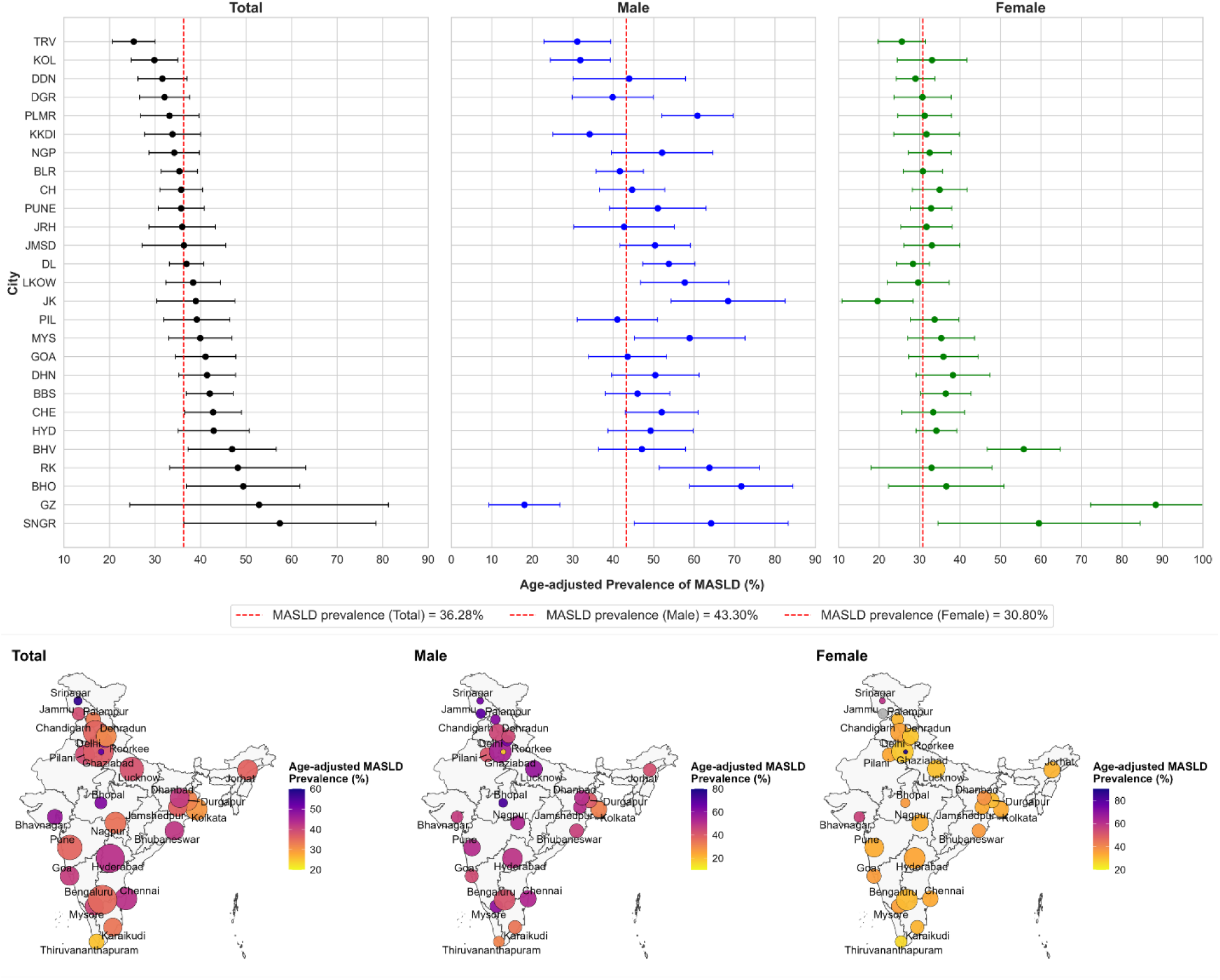
Prevalence of age-adjusted MASLD in different cities against the total population, gender: Male and Female. The red dotted line indicates the age-adjusted prevalence for the overall group, with the population group in parentheses. The panel below shows similar data in a map of India, with the size of the circle indicating the population screened at that city (Data in Supplementary Table 5). The city is the location of the Sample Collection Institute. Abbreviations: TRV, Thiruvananthapuram; KOL, Kolkata; DGR, Durgapur; DDN, Dehradun; PLMR, Palampur; KKDI, Karaikudi; CH, Chandigarh; PUNE, Pune; JMSD, Jamshedpur; BLR, Bengaluru; JRH, Jorhat; DL, Delhi; LKOW, Lucknow; MYS, Mysore; NGP, Nagpur; PIL, Pilani; RK, Roorkee; JK, Jammu; GOA, Goa; DHN, Dhanbad; BBS, Bhubaneswar; HYD, Hyderabad; CHE, Chennai; BHV, Bhavnagar; BHO, Bhopal; GZ, Ghaziabad; SNGR, Srinagar.

### Prevalence of cardiometabolic risk factors (CRMFs) across different subgroups in MASLD

The burden of cardiometabolic risk factors (CRMFs) in MASLD was substantial, with ∼32% of individuals carrying three CRMFs, 24% carrying four, and 14% carrying all five (Figure 3). Risk factor clustering rose steeply with age, with the prevalence of all five CRMFs reaching 24% among those ≥60 years. Geographic variation was also evident (Supplementary Figure 2): cities such as Jamshedpur, Dhanbad, Goa, Karaikudi, and Bhopal showed the highest clustering of ≥4 CRMFs (>20%), whereas Thiruvananthapuram, where MASLD prevalence was also lowest, had the fewest individuals with all five CRMFs (<10%). In contrast, without-MASLD participants exhibited a much more favourable risk profile, with the majority having only one or two CRMFs and <6% carrying all five. Taken together, these findings highlight both the age-dependent accumulation of cardiometabolic risk in MASLD and regional pockets of particularly high clustering, suggesting interaction between biological ageing, urbanization, and local lifestyle factors.

**Figure 3:**
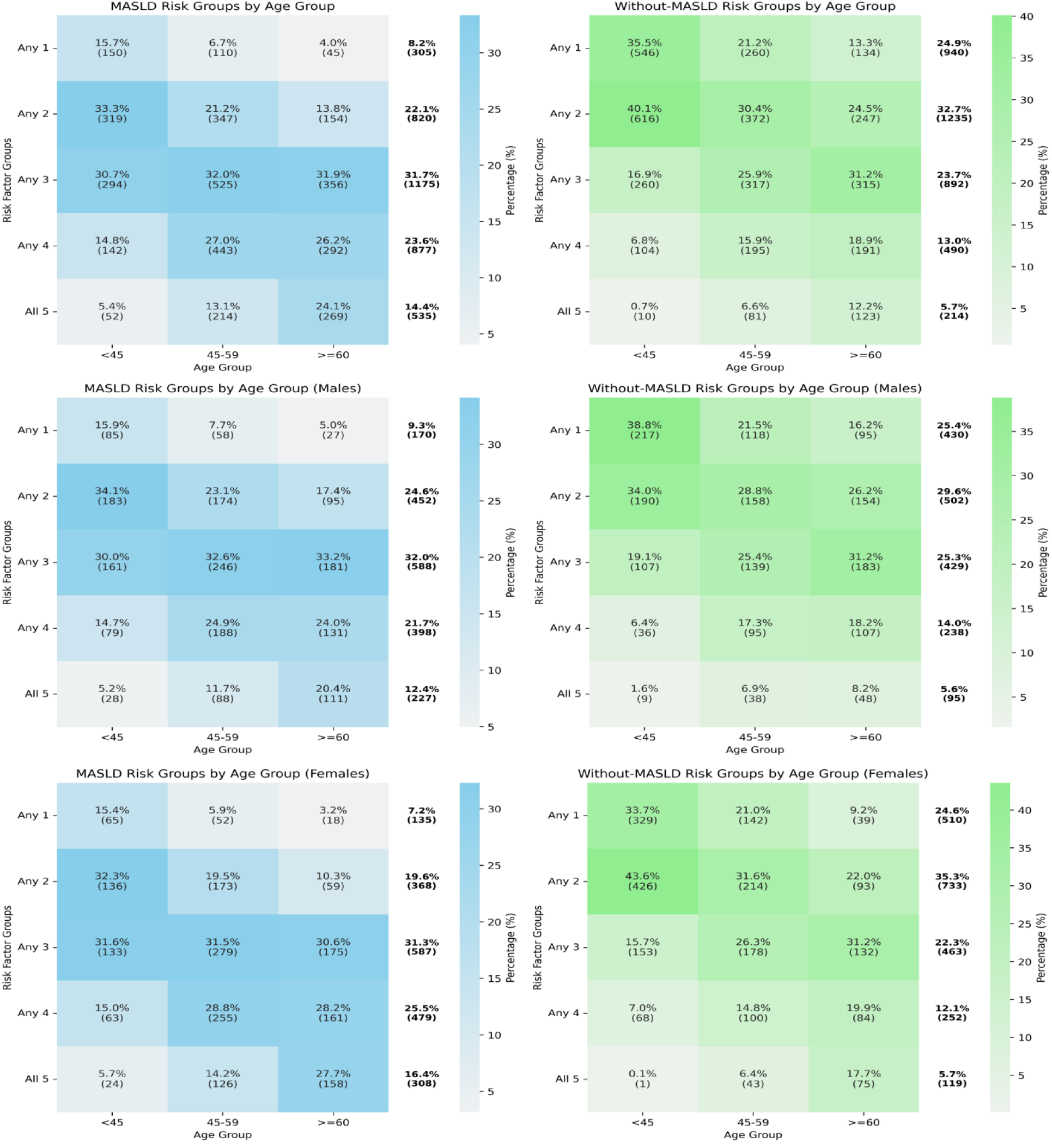
Stratification of CMRF in MASLD population by age and gender within MASLD (blue) and without MASLD groups (green). Data are presented as percentages within age groups of CMRFs (vertical). Outside figure percentages and numbers reflect the total MASLD and the without-MASLD group contribution (horizontal). Without-MASLD may not add to the completeness, as other participants had zero risk factors (281 in total, 142 in males and 139 in females).

### Prevalence of liver fibrosis in the cohort

Among 7,715 participants in total, crude prevalence of liver fibrosis (≥F2) was 3.9%, (F2-1.8%, F3-1.4%, F4-0.7%) while in 3688 with MASLD where LSM data was available, the rate was 6.3% (F2-2.9%, F3-2.4%, F4-1.0%) respectively with consistently higher rates in men across all stages (Supplementary Figure 3A). Fibrosis prevalence increased progressively with age, particularly for the F2 category, while cirrhosis (F4) was largely confined to individuals over 60 years (Supplementary Figure 3B-D), consistent with the late onset of progression to end-stage diseases. Age-adjusted fibrosis was 2.3% overall and 2.5% and 2.4% in males and females, respectively overall.

Stratification by BMI and glycemic status revealed markedly higher crude fibrosis burden in diabetes and obesity. The prevalence of F2 fibrosis was notably elevated among individuals with diabetes (4.02%), followed by those with prediabetes (1.81%), and individuals in obesity categories I and II (4.89%). In contrast, the prevalence of F3 fibrosis was highest in the obesity II category. The prevalence of cirrhosis (F4) approached 2% in both diabetic and obesity II groups (Supplementary Figure 4A-B).

Geographic variation was pronounced, with the highest age-adjusted prevalence of significant fibrosis (≥F2) observed in Jorhat (10.2%), Bhopal (4.7%), Jammu (4.39%), Delhi (4.19%), Bhubaneswar, and Roorkee, and the lowest in Kolkata (0.26%) (Figure 5, Supplementary Table 6). Delhi and Jammu had the highest age-adjusted prevalence of ∼1% Cirrhosis (Figure 4). These findings highlight the dual influence of age and metabolic risk on fibrosis progression in MASLD, alongside substantial regional heterogeneity.

**Figure 4:**
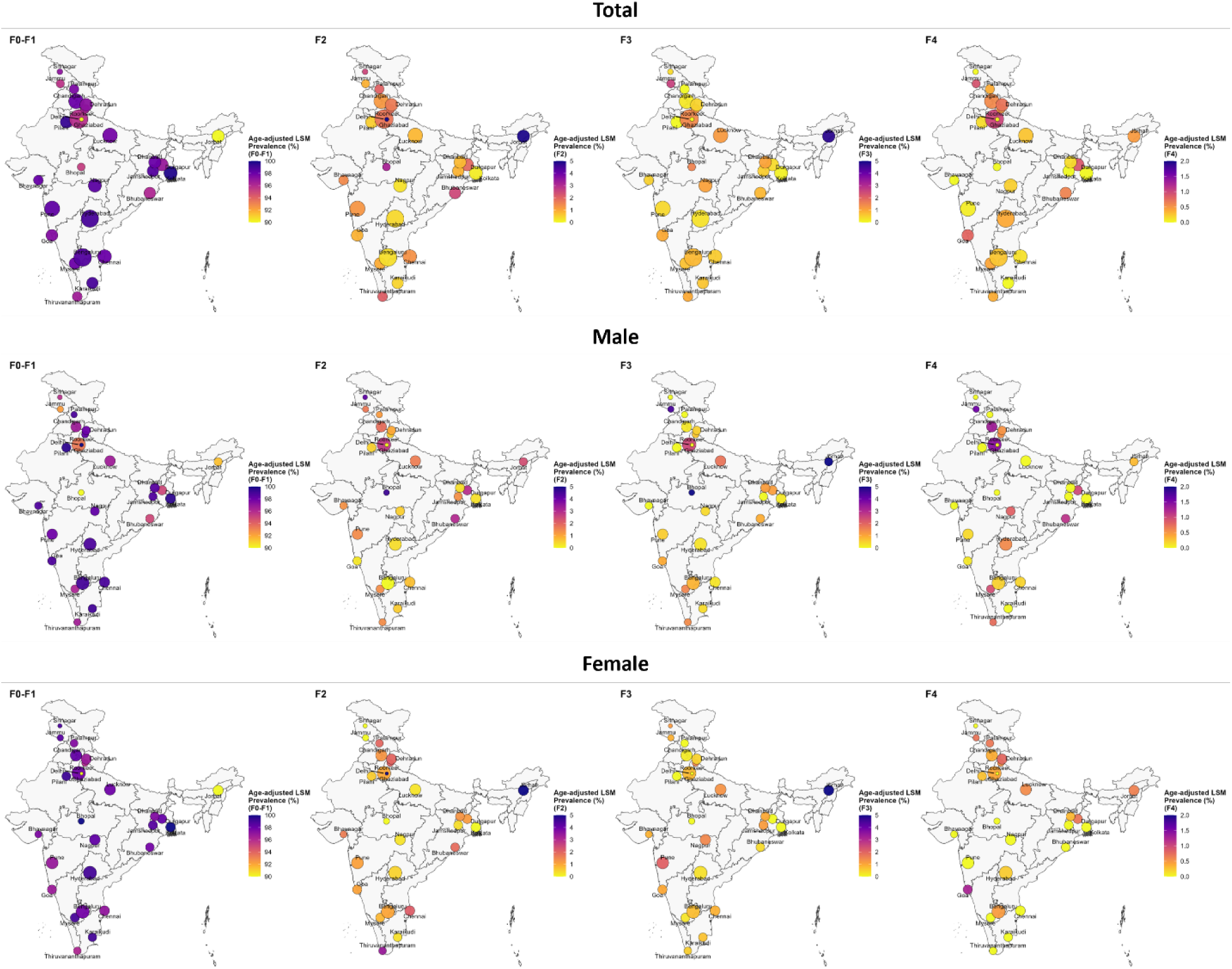
Prevalence of age-adjusted Fibrosis as per LSM grades in different cities against the total population (n=7715), gender: Male and Female. The panel shows data in a map of India, with the size of the circle indicating the population screened at that city (Data in Supplementary Table 6). The city is the location of the Sample Collection Institute.

**Figure 5:**
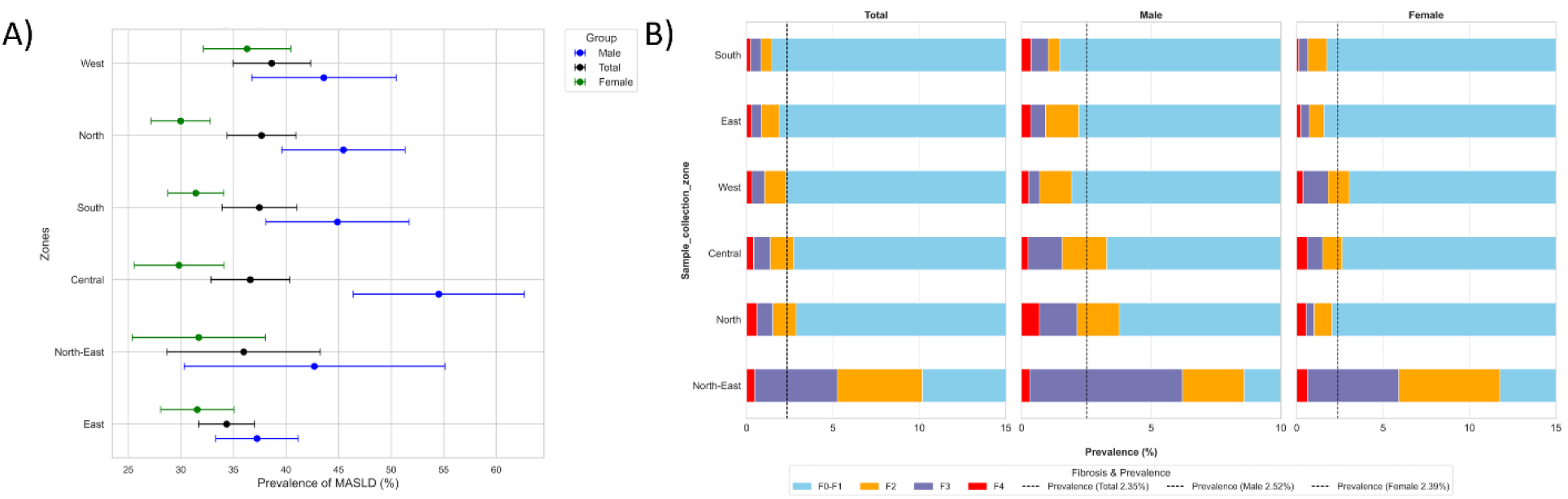
Zone-wise age-adjusted prevalence of MASLD (A) and normal F0-F1 score with F2, F3, and F4 fibrosis (B) in Phenome India Cohort. The dotted line indicates the age-adjusted prevalence for the overall group, with the value in parentheses in fibrosis (Data in Supplementary Tables 8 and 9).

### Regression Modelling for Liver Fibrosis

Multivariate regression identified age, diabetes, obesity, and central adiposity as key determinants of liver fibrosis. Compared with individuals aged 20–30 years, the risk of fibrosis rose progressively with age, becoming significant beyond 60 years (OR 4.77, 95% CI 1.1–20.70 for 60–70 years; OR 4.90, 95% CI 1.2–20.0 for ≥70 years). Diabetes was one of the strongest independent predictors (OR 3.12, 95% CI 2.30–4.0, p<0.001), whereas hypertension and dyslipidemia were not associated. Among BMI categories, Obese II conferred a markedly elevated risk (OR 4.2, 95% CI 1.9–9.5, p<0.001), while Obese I showed only a borderline effect in terms of significance. In addition, an elevated waist–hip ratio was independently associated with a two-fold higher risk of fibrosis (OR 2.1, 95% CI 1.3–3.5, p<0.01). Together, these findings highlight that fibrosis risk in this cohort is driven primarily by advancing age, diabetes, and central obesity, with the highest risk concentrated in those with severe obesity and abdominal adiposity (Supplementary Table 7). While MASLD seems predominantly obesity-driven with contributions from metabolic comorbidities, progression to fibrosis largely seems to be determined more by age, diabetes, and central adiposity.

### MASLD and Fibrosis Distribution Amongst Different Zones

Regional patterns of MASLD and fibrosis demonstrated both overlap and divergence (Figure 5 A–B, Data in Supplementary Tables 8 and 9). MASLD prevalence was highest in men from the central and northern zones, with women showing higher prevalence in the west, followed by north-east, east and south and lowest in the north and central. Significant fibrosis (≥F2), however, was disproportionately elevated in the north-east, with high MASLD zones such as the north and central also showing correspondingly high fibrosis rates. This discordance, particularly the excess fibrosis in the north-east, suggests that additional contributors beyond MASLD may be driving liver injury. These observations raise the possibility that without-MASLD fibrosis may represent an important, under-recognised burden in specific regions (Figure 4B).

### Assessment of Fibrosis in participants with and without-MASLD

Among individuals with liver fibrosis, nearly 29% did not have MASLD, highlighting a substantial subgroup that would ordinarily not have been tested for fibrosis. Compared with MASLD fibrosis, those without-MASLD were leaner, with significantly lower weight (74.7 vs. 65.1 kg, p<0.01), BMI (29.6 vs. 25.1 kg/m², p<0.01), waist–hip ratio (0.94 vs. 0.91, p<0.01), and CAP scores (307.5 vs. 216 dB/m, p<0.01). They were more often in the normal or overweight BMI range (11.1% vs. 47.8%) and far less likely to fall in Obese II (45.3% vs. 11.6%). In line with this anthropometric profile, they exhibited fewer metabolic abnormalities, including lower ALT (38.3 vs. 26.7 U/L, p<0.01), fasting glucose (112.1 vs. 97.8 mg/dL, p<0.01), HbA1c (6.5% vs. 5.9%, p<0.01), triglycerides (126.8 vs. 94.6 mg/dL, p<0.01), LDL cholesterol (109.3 vs. 94.0 mg/dL, p<0.01), HOMA-IR (4.22 vs. 1.98, p<0.01). The prevalence of dyslipidemia (56% vs. 30.4%) and diabetes (62.0% vs. 43.5%) was also significantly lower, while age, sex distribution, hypertension, and menopausal status did not differ. Despite this more favourable metabolic and anthropometric profile, these individuals had comparable fibrosis stage distribution (F2– F4) to MASLD fibrosis. This striking discordance suggests that nearly one-third of fibrosis cases may arise through pathways independent of obesity and classical metabolic risk, raising the possibility that inflammation or other non-traditional mechanisms contribute to disease in this group (Table 4 and Supplementary Table 9).

**Table 4:**
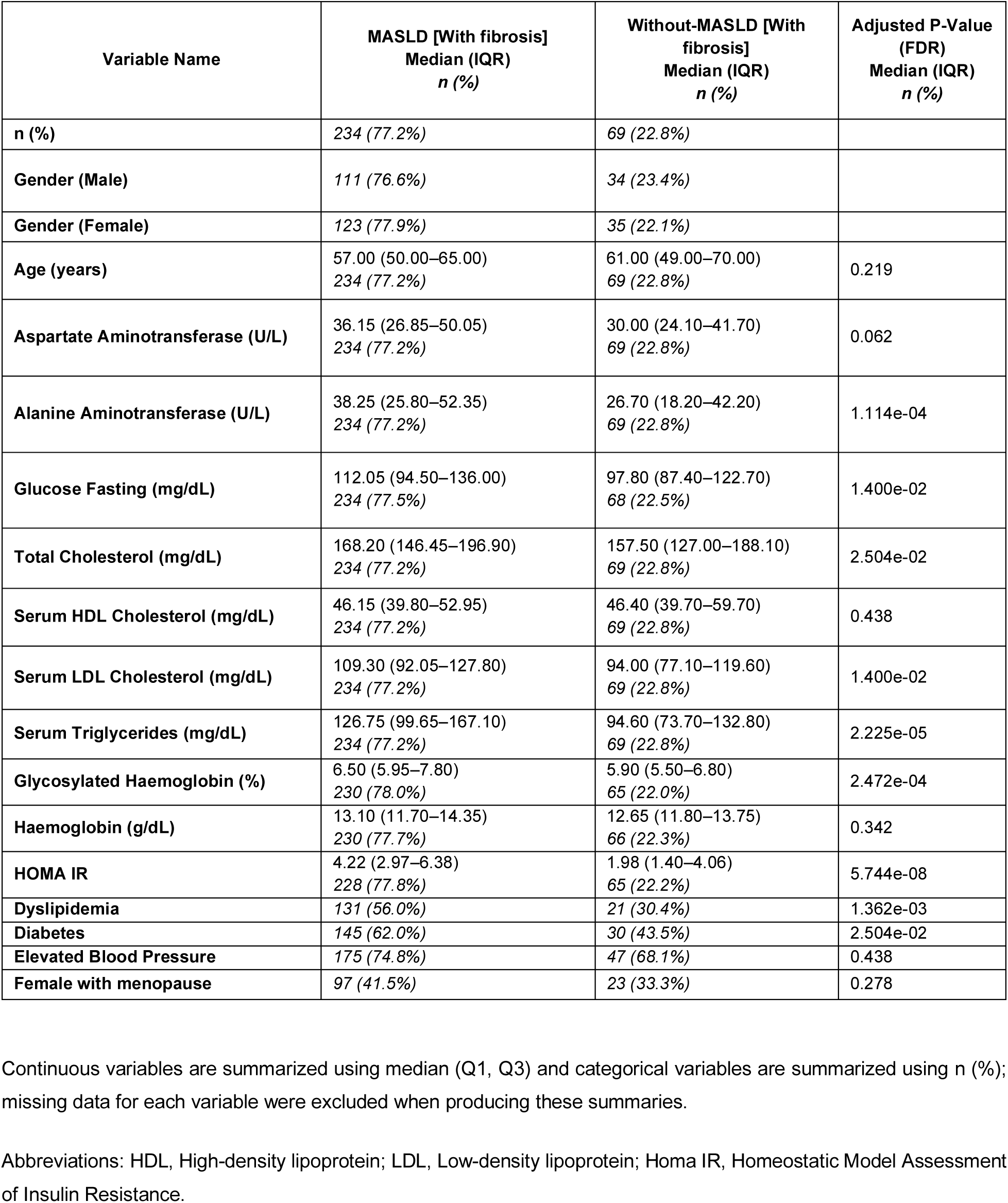
Fibrosis without and with MASLD.

| Variable Name | MASLD [With fibrosis]<br>Median (IQR)<br>n (%) | Without-MASLD [With<br>fibrosis]<br>Median (IQR)<br>n (%) | Adjusted P-Value<br>(FDR)<br>Median (IQR)<br>n (%) |
| --- | --- | --- | --- |
| n (%) | 234 (77.2%) | 69 (22.8%) |  |
| Gender (Male) | 111 (76.6%) | 34 (23.4%) |  |
| Gender (Female) | 123 (77.9%) | 35 (22.1%) |  |
| Age (years) | 57.00 (50.00–65.00)<br>234 (77.2%) | 61.00 (49.00–70.00)<br>69 (22.8%) | 0.219 |
| Aspartate Aminotransferase (U/L) | 36.15 (26.85–50.05)<br>234 (77.2%) | 30.00 (24.10–41.70)<br>69 (22.8%) | 0.062 |
| Alanine Aminotransferase (U/L) | 38.25 (25.80–52.35)<br>234 (77.2%) | 26.70 (18.20–42.20)<br>69 (22.8%) | 1.114e-04 |
| Glucose Fasting (mg/dL) | 112.05 (94.50–136.00)<br>234 (77.5%) | 97.80 (87.40–122.70)<br>68 (22.5%) | 1.400e-02 |
| Total Cholesterol (mg/dL) | 168.20 (146.45–196.90)<br>234 (77.2%) | 157.50 (127.00–188.10)<br>69 (22.8%) | 2.504e-02 |
| Serum HDL Cholesterol (mg/dL) | 46.15 (39.80–52.95)<br>234 (77.2%) | 46.40 (39.70–59.70)<br>69 (22.8%) | 0.438 |
| Serum LDL Cholesterol (mg/dL) | 109.30 (92.05–127.80)<br>234 (77.2%) | 94.00 (77.10–119.60)<br>69 (22.8%) | 1.400e-02 |
| Serum Triglycerides (mg/dL) | 126.75 (99.65–167.10)<br>234 (77.2%) | 94.60 (73.70–132.80)<br>69 (22.8%) | 2.225e-05 |
| Glycosylated Haemoglobin (%) | 6.50 (5.95–7.80)<br>230 (78.0%) | 5.90 (5.50–6.80)<br>65 (22.0%) | 2.472e-04 |
| Haemoglobin (g/dL) | 13.10 (11.70–14.35)<br>230 (77.7%) | 12.65 (11.80–13.75)<br>66 (22.3%) | 0.342 |
| HOMA IR | 4.22 (2.97–6.38)<br>228 (77.8%) | 1.98 (1.40–4.06)<br>65 (22.2%) | 5.744e-08 |
| Dyslipidemia | 131 (56.0%) | 21 (30.4%) | 1.362e-03 |
| Diabetes | 145 (62.0%) | 30 (43.5%) | 2.504e-02 |
| Elevated Blood Pressure | 175 (74.8%) | 47 (68.1%) | 0.438 |
| Female with menopause | 97 (41.5%) | 23 (33.3%) | 0.278 |

Continuous variables are summarized using median (Q1, Q3) and categorical variables are summarized using n (%); missing data for each variable were excluded when producing these summaries.
Abbreviations: HDL, High-density lipoprotein; LDL, Low-density lipoprotein; Homa IR, Homeostatic Model Assessment of Insulin Resistance.

### Cytokines Analysis in MASLD vs without-MASLD sub-group with and without fibrosis

Growing evidence also indicates that an inflammatory milieu plays a critical role in the development and progression of MASLD, through low-grade, systemic chronic inflammation (29). Therefore, to dissect the contribution of systemic immune mediators, we profiled 48 cytokines, chemokines, and growth factors using a multiplex assay across patients stratified into four clinical subgroups based on the presence or absence of MASLD and liver fibrosis.

Across the cohort, twenty-five of 48 cytokines were found to be significantly different: Eotaxin, FGF basic, G-CSF, HGF, IFN-γ, IL-1a, IL-1b, IL-1ra, IL-2Ra, IL-4, IL-6, IL-7, IL-8, IL-9, IL-16, IL-17A, IL-18, IP-10, M-CSF, MCP-1(MCAF), MIP-1a, MIP-1b, RANTES, SCF, and TRAIL, all of which were higher in the MASLD group vs the without-MASLD group, irrespective of fibrosis status (Supplementary Table 11). When restricting the analysis to individuals without fibrosis, 16 of 48 analytes were significantly elevated in the MASLD group. These comprised predominantly pro-inflammatory mediators, including cytokines (IL-1α, IL-1β, IL-7, IL-8, IL-16, IL-18), chemokines (CCL11 (Eotaxin), CXCL10 (IP-10), CCL2 (MCP-1), CCL3 (MIP-1a), TRAIL), and growth factors (G-CSF, HGF, M-CSF), consistent with a systemic inflammation profile in MASLD (Supplementary Table 12, columns 2-4).

Comparison of MASLD participants with or without fibrosis revealed a striking amplification of this inflammatory signature. All 16 cytokines and chemokines elevated in MASLD alone were further increased in the fibrosis sub-group. Molecules associated with type 1 immune responses, such as IFN-γ, IL-12, and CXCL10 (IP-10), were approximately twofold higher in the fibrosis sub-group, suggesting that the transition to fibrosis is accompanied by an intensified Th1-skewed inflammatory milieu (Supplementary Table 13).

To distinguish whether these changes were primarily driven by MASLD or by fibrosis per se, we next examined individuals with fibrosis but without clinical MASLD. Interestingly, the same inflammatory mediators elevated in MASLD fibrosis were also significantly increased in fibrosis without-MASLD, indicating that fibrosis, rather than being driven by steatosis, may have a contribution from systemic inflammation. Moreover, compared to MASLD with fibrosis, the fibrosis-associated without-MASLD sub-group exhibited further elevation of 11 additional inflammatory mediators (β-NGF, IL-2, IL-3, IL-12p70, IL-12p40, IL-13, IL-15, IL-17A, CXCL9 (MIG), CCL5 (RANTES), CXCL12 (SDF-1a)), highlighting a broader pro-inflammatory network specifically enriched in fibrosis independent of MASLD (Supplementary Table 12, columns 5-7).

## Discussion

Our nationwide analysis reveals a substantial MASLD burden, with an overall crude prevalence of 47.8% and an age-adjusted prevalence of 36.3%. While no major studies are available from India for population-wide prevalence, a previous study for NAFLD from India showed a pooled prevalence of ∼38 % in adults, and similarly, globally was shown to be 38% (30). Our study showed that prevalence had regional variance, being higher in the western, central, and northern zones in India.

NAFLD is the most prevalent chronic liver disease worldwide (31), with the highest prevalence reported in Latin America (44%), the Middle East and North Africa (36.5%), South Asia (33.8%) followed by North America (31%) and Western Europe (25%) (31). While there are no major countrywide studies in India for the community-based prevalence of MASLD in the general population, India’s pooled NAFLD prevalence is ∼38.6% (95% CI 32-45.5) (2), and high urban estimates (∼56.4% MAFLD in Delhi and ∼61.5% NAFLD in Chennai cohorts) mirror this trend (5, 32). Importantly, 6.3% individuals with MASLD were found to have significant liver fibrosis. These figures highlight the subclinical nature of early MASLD and its silent progression, which poses a potential burden to the public health systems if left unaddressed.

The strong association of MASLD and fibrosis with metabolic comorbidities, IR and HTN emphasizes the need for integrated management of these comorbidities to halt disease progression and prevent both liver-related and non-liver-related events in these patients.

Notably, in our study, 66.4% participants with Diabetes had MASLD (1280 of 1927), while 9.1% (175 of 1915) had significant fibrosis. Recently, a clinic-based retrospective cohort study reported any degree of steatosis and fibrosis in 75.6% and 28.6% of 1070 patients with T2DM (33). In the “MAP” study, which was again a clinic-based retrospective study, the prevalence of MASLD and fibrosis was reported to be 68.2% and 33.7% respectively. However, it is to be noted that the MAP study used a lower cutoff for both CAP (38) and F2 (7-10 kPa) (15). Among overweight and obese individuals, fibrosis prevalence was high (14.25%).

In a recently published review, Hagström, H et al. reported that in the last 2 years, they have observed MASLD prevalence in the general population to be 38% while in the Diabetic population, it is 65% in adults. We also observed 36.3% age-adjusted prevalence of MASLD and 66.4% crude prevalence of MASLD in Diabetics, which are similar to what has been stated for properly conducted prevalence studies (14). While we observed relatively lower numbers for fibrosis in our study, likely, this could be due to the fact that ours was community-based rather than clinic-based sampling, unlike the MAP study (15).

As reported previously from an urban population of 6146 individuals from Delhi by Prabhakar T et al, males and females appear to be equally affected by MAFLD with an overall prevalence of approximately 56%, while in our study, we observed the prevalence of MASLD to be 45.85% in females and 49.98 % in males (5). We also observed a higher prevalence of MASLD with increasing age (maximum at the 50-59 age group), similar to the study by Prabhakar T et al (5).

Among various BMI subgroups, we had a similar number for lean MASLD as other published literature for prevalence in the Asian population (34, 35). Our data also confirm the presence of lean MASLD, particularly associated with T2DM, likely reflecting greater visceral rather than subcutaneous adiposity. This suggests poorer glycaemic and lipemic control in individuals with lean MASLD, likely attributed to a greater quantity of visceral fat (rather than subcutaneous), and needs to be explored further. A significant number of post-menopausal females had obesity related MASLD, which is likely because of adiposity distribution pattern changes post menopause (36, 37).

The higher prevalence of fibrosis in individuals with MASLD as compared to those without MASLD, particularly among young and middle-aged individuals, underscores the need for targeted screening and preventive interventions. Fibrosis in MASLD is predominantly driven by IR and dysglycemia. This metabolic milieu promotes lipotoxicity and oxidative stress, accelerating hepatic stellate cell activation and extracellular matrix deposition, which are central to fibrosis progression (38, 39).

Our data suggests a notable regional disparity, with higher prevalence rates concentrated in northeastern (Jorhat) and northern (Delhi, Jammu) and central zones (Bhopal), suggesting contributions from specific regional factors such as dietary, genetic, or environmental influences. Lower prevalence in southern and eastern zones may reflect differences in lifestyle, healthcare access, or screening practices. The regional disparity was also reported in the “MAP” study (15). Similar to our observations, the MAP study also reported a lower prevalence of fibrosis in eastern and southern cities; however, unlike our study, they found a low prevalence of fibrosis in the North-East. This could potentially be due to selection bias in hospital-based studies and the need for further community-based studies to understand the true prevalence of MASLD in our country. This geographic variation highlights the importance of tailored public health strategies to screen and identify liver fibrosis early, particularly in high-prevalence areas, and highlights the need for further investigation into region-specific etiological factors (15).

Cytokines are critical drivers of liver fibrosis in mediating inflammation and fibrogenesis. In MASLD, pro-inflammatory cytokines (TNF-α, IL-6, and IL-1β) fuel chronic inflammation, progressing from simple steatosis to MASH and fibrosis (40–43). Our study revealed elevated levels of both pro-(IFN-γ, IL-1β, IL-8, IL-18, IP-10, MCP-1, MIP-1α), and anti-inflammatory cytokines in MASLD, suggesting a complex interplay of inflammatory and regulatory responses. Without biopsy data, tissue-specific insights were limited. The strong upregulation of Th1-associated cytokines such as IFN-γ, IL-12, and CXCL10 underscores the role of type 1 immune responses in fibrotic progression. From a translational perspective, these results imply that cytokine signatures could serve as biomarkers to distinguish MASLD with or without fibrosis, and potentially fibrosis of other etiologies.

Our findings advocate for policy shifts to include TE-based screening in primary healthcare centres, particularly in urban and high-risk rural areas. Training healthcare workers to use TE and integrating it with existing metabolic disease programs could enhance scalability.

There are some limitations in our study, which include reliance on TE rather than histology, the exclusion of individuals with any alcohol intake, and the urban/semi-urban bias of the cohort. Nonetheless, this is the largest community-based study of MASLD in India to date, and its integration of epidemiology, metabolic risk factors, and immunological profiling provides a comprehensive understanding of disease burden and biology. Our findings underscore the need for early detection, integration of liver health into metabolic disease programs, and translational research into cytokine-driven pathways as therapeutic targets.

## Conclusion

The high prevalence of MASLD (47.81%) and associated fibrosis (∼7%) in India represents an urgent public health challenge. Future longitudinal studies are needed to track MASLD and fibrosis trends and evaluate the impact of screening interventions.

## Ethics

The study was approved by IHEC at IGIB (reference no.: CSIR-IGIB/IHEC/2023-24/16). The study was registered vide CTRI number-CTRI/2024/01/061807.

## Acknowledgments

We acknowledge the support from CSIR, participants, and volunteers. Further, the following names are acknowledged:

Arpana Parihar, Shalu Yadav, Mohd Abubakar Sadique, Akshay Singh Tomar, Varsha Agrawal, Sakshi Rajput, Aabha Kushwah, Shilpee Chauhan, Varsha Parmar, Pushpesh Ranjan, Neeraj Kumar, Abhisek Giri, Navnidhi Tripathi, Jyoti Lodhi, Parul Shrivastava, Prachi Shrivastava, Komal Sharma (**CSIR-AMPRI**), Surya Narayan Mishra, Sathya Swetha, Gururaj Kalshetti, Rakesh Pal Bhagat, Chandreshwar Raju Kataru, Hari Charan Goud Nerella, Amit Chakraborty, Prangya Paramita Sahoo, Alagu Sankareswaran, Sofia Banu, Priyadarshini Sanyal, Sourav Ganguly, Manash Kumar Behera, Pallavi Rao T, Akila Ramesh, Niharika Tiwary, Devika Mahimakar, Abhishek Saha, Shreya, Pramoda, Ruqaiya Tasneem (**CSIR-CCMB**), Sangavi Pakkyam (**CSIR-CECRI**), N Vinodkumar, Rani C Avilash Sumithra, Pramila Epparti, M Archana (**CSIR-CFTRI**), Debabrata Chanda, Dnyaneshwar Umrao Bawankule, Pankhuri Singh, Pankaj Kumar Shukla, Parmanand Kumar, Sumati Sen, Chandra Kant, Sarita Upadhyay, Sumit Kushwaha, Sukriti Srivastava, Kavita Singh, Om Prakash Choudhary Kar, Debasmita Sahoo, Anant Kumar Chaudhary (**CSIR-CIMAP**), Srinivetha Pathmanapan, Syed Nasar Rahaman, Vikash Negi, Jegan K, Sankar Palanivel, Lishadevi M, Pranathy K, Kirubakaran BJ, Thiyaneshwar (**CSIR-CLRI**), Sujata Pachhal, Sukanta Maji, Sonali Garai (**CSIR-CMERI**), Darshan Singh, Mukesh Kumar, Sakshi Gupta, Keshav Kaushik, Satyajit Nayak, Lakshmy T (**CSIR-CRRI**), Sandeep Kumar, Rajni, Preetismita Borah, Manish Kumar, Goraj Singh, Suraj Prakash, Ajay Kumar, Inderjit, Shubham Patial, Shavita, Amandeep, Jyoti Jangra, Aman Grewal, Shivalika, Rajrani, Swapna, Dinesh Kumar, Dhiru, Munish, Akriti, Deepak Kumar, Gourav, Komal, Prashant Kumar, Amrit Marthu, Rishav Rai, Amir Khan, Satinder Kaur, Netra, Pankaj, Anuj Sharma, Parneet Kaur, Krishna, Pallavi Suman, Abhishek, Yash (**CSIR-CSIO**), Salwa Naushin, Subhash Gurjar, Rajesh Kumar, Anushka (**CSIR-IGIB**), Anuj, Aditya Ranout, Trilok Saini, Sahdev Choudhary, Amit Kumar, Shweta Sharma, Neha Bhardwaj, Kajal Kalia, Suresh Kumar, Abhishek Goel, Ravi Kumar, Surbhi Mali, Komal Goel, Swati Katoch, Shagun Dogra, Vinesh Sharma, Rajneesh Kumar, Poonam Dhiman, Pardeep Poonia, Sahiba Chahal, Rahul Kumar, Athrinandan S. Hegde, Rupinder Kaur, Vivek Kumar, Prakriti Sharma, Avisha Sharma, Ankita, Mohak Mali, Priyanka Koundal, Ashruti (**CSIR-IHBT**), Rajendra Prasad Punna, Kiran Kumar A, Taslim B Shaikh, Nidhi Sharma, Anjali Veeram, Komal walvekar, Mani Sharma, Suriya panneer, Rajwinder Kaur, Ramasatyasri Kotipalli, Hari Priya Sripadi, Vaishnavi Kambhampati, Pinkal Sondarva, Divya Vani kundurthi, Maneesha Yeddula, Sindoora Dhoolipala (**CSIR-IICT**), Harleen Kaur, Ayan Banerjee, Rahul Gautam, Mirnalini Juyal, Mridul Budakoti, Preetanshika Tracy, Pratima Ashoke Patel, Ritu Mourya, Harish Panwar (**CSIR-IIP**), Sneha Mohanty, Mohd Tauseef Khantwal, Vaibhavi Lahane, Shreya Tripathi, Sunita Devi, Gulafsha Siddiqui, Km Priya, Priyanka Goswami, Sandip Chatterjee, Sakshi Singh, Sachin Mishra, Abhineet Singh, Vishal Narayan Soni, Devendri Khantwal, Murk Kumari, Sanjivani Naik, Apoorva Dewangan, Ananta Joshi, Akansha Chaurashiya, Akansha Chauhan, Sheetal Agarwal, Joel Saji, Nikita Shraogi (**CSIR-IITR**), Ankita Das, Mayuri Kumari, Amit Kumar Swain, Deepak Kumar Ahirwar, Nilotpal Kapri, Ghrutanjali Sahu, Santosh Kumar Behera, Suchismita Senapati, Subhashree Nayak, Yogesh Chaudhary, Anagonou Bertrand, Bibekananda Nayak, Suraj Kumar Sahu, Saswati Suryasnata, Champakeswara Mahanta, Annapurna Behera, Satyabrata Sahoo, Akshyurna Pattnaik, Rakhi Rani Biswas, Chandrama Dipa, Adyasha Bijay Mishra, Dinesh Kumar Panda, Sudipta Nayak, Avishek Kar, Kajal Sundaray, Swagatika Dash, Alok Pattanaik, Sonali Swapanjali Bhoi, Swikruti Mishra, Shalini Das, Parinita Mishra, Debidatta Barik, Tejaswini Das, Subhashree Pattnaik (**CSIR-IMMT**), Kailash T Bhamare, Chander Shekhar Sharma, Amit Kumar1, Vineet Kumar, Deepak Bhatt, Surjeet Singh, Harminder Singh, Paramjit Lal, Sandeep Kumar, Nitin Sharma, Jaideep Mehta, Renu, Rohtas Ranga, RK Dhiman, Anjali Koundal, Bhumika Vaidya, Amit Kumar2, Shweta Pandey, Anunay Sinha, Shubham, Bulbul Roy, Pompi Bhadra, Meena Sharma, Sana Khatun, Payal Thakur, Bhagyashree Rabha, Meenal Rastogi, Mayur Sudhakar Zarkar, Somnath Chindhe, Shreya Singh, Neelam, Rakesh Kumar, Siddhakam Palmal, Lalit Kumar, Joyasree Das, Parvez Ahmad (**CSIR-IMTECH**), Kiran Raj S, Ravi Kumar Sharma Kumar (**IPU, CSIR Hqrs.**), Nataraj D, Swetha C Desai, Shivanna TB, Murthy BYK, Raghu G, Sunil A, Shashidhara KN, Raghavendra Swamy M, Aravind Kumar Joseph, Purushotham HM, Shiva Kumar SR, Muniprakash M, Shekhar Gogeri, Siva Kumar R, Jaikesh S, Ranganatha S, SatishKumar Shri, Shashikala U, Praveen Kumar JD, Jitendra Ping (**CSIR-NAL**), Nikhilesh Yadav, Parveen Goyal, NMR Ashwin, Shashikala Ranjane, Abujunaid Khan, Priyanka M Bankar, Rachel Samson, Ashish S Jagtap, Amreen Sheikh, Shahaji Palaskar, Jugal Kanerkar, Arvind Chourasiya, Bhagyashree Likhitkar, Pradeep S, Deepak Jadhav, Pranay Awathare, Swaraj Jathar, Pooja Deshmukh, Vaishnavi Salunkhe, Ashtamy MG, Yugendra Patil, Rohit S Dashpute, Minal R Bhalerao, Vinod Kamble, Yashodhara Shinde, Vineetkumar S Nair, Prathmesh Ghongade, Prashant Kalaskar, Ganesh Jadhav, Sakshi Pingle, Preshitta Bhat, Shubham Choure, Tanaji B Devkate, Vikram Nichit, Sonali Gaikwad, Abhishek Tripathi, Sayli Jamdade, Shubham Kumar, Rutuja Ugle, Kaumudi Joshi, Khushboo Bavishi, Priya Bhavsar, Nandini Rangari, Ayushi Rushesari, Maria Kuwazerwala, Taha Farhan Siddiqui, Anand Kumar Shukla, Prakashini Saroj Nilgirwar, Sakshi R Mangate, Sindhu Bali, Prabha Oli, Ameya A Pawar, Rashdajabeen Q Shaikh, Ankita Namdeo, Disha Dwivedi, Suryaprakash (**CSIR-NCL**), Pankaj Pardhi (**CSIR-NEERI**), Jayashree Chiring Phukon, Shridhar Hiremath, Prachurjya Dutta, Parishmita Borgohain, Moirangthem Goutam Singh, Devpratim Koch, Dipanneeta Das Gupta, Pankaj Barman, Sahana SK, Monojit Kumar Roy, Aditya Sarkar, Bhaben Sharma, Shyamalima Mech, Masum Saikia, Trishna Rani Borah, Gayatri Gogoi, Anupriya Borah, Udeshna Changmai, Himadri Das, Anshuman Goswami, Rocktotpal Mahanta, Rina Yumnam, Sukanya Borthakur, Manabendra Borah, Sarangthem Dinamani Singh, Pronami Gogoi, Priyanka Saikia, Umakanta Tanti, Mohan Kurmi, Ravi Kumar Sahu, Nayan Jyoti Borah, Babli Borah, Sindhu Sharma, Trishna Dutta, Sarifa Mafuz Ahmed, Bidyut Prakash Deka, Darshana Tamuli, Meghna Kakoty, Bhaswati V. Borah, Yashodhara Goswami, Priyanka Boro (**CSIR-NEIST**), Inshamol KP, Athulya, Akhila, Amrutha M, Anaga, Anaswara P A, Anirudh, Anjitha, Anupama, Anusha, Anusree P, Aparna, Aparna Dinil, Arathy, Archa R S, Archana, Arnold, Athira V C, Biji Raphy, Dileep R Nair, Evan, Gayathri, Gopika R, Greeshma, Greeshma Jayan, Karthika N, Karthika Nath, Kavya Mohan, Krishan Unni, Lakshmi M Nair, Lakshmi Shaji, Nandana A B, Neeraja, Neeraja M, Neetha P Sobandas, Parvathy, Reena R, Saranyadevi, Shahansha, Shamna Fathima, Shehbas C, Shilpa, Sruthi, Swapna B, Varsha, Vishnu, Valan Rebinro, Adarsh VP, Jedy Jose (**CSIR-NIIST**), Govind Ranade, AS Unnikrishnan, Sohan Pal Meena, Karishma Pradeep Chari, Vitasta Jad, Vikash Kumar, Nishamol M, Helen Agnes, Natasha Maria Barnes, Bhumika Rohidas Shirodkar, Shania Waluscha Moeres, Yuvrani Halarnkar, Ramila Ram Gaonkar, Mrinalini Chandra Mohan, Pratibha Bachhley (**CSIR-NIO**), Himani Meena, Abhinav Banait, Shekar Sharma (**CSIR-NIScPR**), Rahul Tirkey, Mousumi Sarkar, Sudip Kumar, Vinod (**CSIR-NML**), Rajesh, Sumana Gajjala, Sudesh Yadav, Vinod Kumar Tanwar, Vishesh Garg, Manoj Kumar Pandey, Vikash Sharma (**CSIR-NPL**).

## Author Contributions

Phenome India Consortium Authors

## Study Planning and Conceptualization

Viren Sardana, Shantanu Sengupta, Kumardeep Chaudhary (**CSIR-IGIB**), Partha Chakraborty (**CSIR-IICB**)

## Project conceptualization

Shantanu Sengupta, Debasis Dash, Viren Sardana, Kumardeep Chaudhary (**CSIR-IGIB**), Giriraj Ratan Chandak, Swasti Raychaudhuri, Karthik Bharadwaj Tallapaka (**CSIR-CCMB**), Mahesh J Kulkarni (**CSIR-NCL**), Partha Chakraborty, Dipyaman Ganguly (**CSIR-IICB**), Umakanta Subudhi (**CSIR-IMMT**).

## Collection management

Shantanu Sengupta, Debasis Dash, Viren Sardana, Kumardeep Chaudhary, Vamsi K. Yenamandra, Aastha Mishra, Ajay Pratap Singh, Swarnendu Bag, Pankaj Pandey, Ankita Sahu, Komal Jindal, Vivek Junghare, Tarani Mathur, Meghana Arvind, Satyartha Prakash, Yogesh Kumar, Vignesh S Kumar, Deeksha Yadav, Deepak (**CSIR-IGIB**), Swasti Raychaudhuri, Giriraj Ratan Chandak, Karthik Bharadwaj Tallapaka, Rakhesh K V (**CSIR-CCMB**), Ashok P Giri, Narendra Y Kadoo, Mahesh J Kulkarni, Dhanasekaran Shanmugam, Mahesh S Dharne, Syed G Dastager, Chiranjit Chowdhury (**CSIR-NCL**), Partha Chakraborty, Dipyaman Ganguly, Shilpak Chatterjee (**CSIR-IICB**), Umakanta Subudhi, Bhabani S Jena, Trupti Das, Boopathy Ramasamy, T Pavan Kumar (**CSIR-IMMT**), Ramakrishna Sistla, Prabhakar Sripadi, Jagadeshwar Reddy Thota, Ramesh Ummanni, Srinivasa Rao M, Sai Balaji Andugulapati (**CSIR-IICT**), Amit Lahiri, Mrigank Srivastava, Vivek Bhosale (**CSIR-CDRI**), Iranna Gogeri (**CSIR-4PI**), Raju Khan, Narendra Singh (**CSIR-AMPRI**), Rajesh Kumar Verma, Neeraj Jain (**CSIR-CBRI**), Ganesh Venkatachalam, Murugan Veerapandian (**CSIR-CECRI**), Deepak Bansal, Amit Kumar, Dinesh Gupta, Dheeraj Kumar Kharbanda, Sk. Masiul Islam (**CSIR-CEERI**), Prakash M Halami, S P Muthukumar (**CSIR-CFTRI**), Anirban Pal, Anil Kumar Maurya (**CSIR-CIMAP**), Jai Krishna Pandey, Bhanu Pandey, A K Raman (**CSIR-CIMFR**), Suresh Kumar Anandasadagopan (**CSIR-CLRI**), Swati Saha, Vishal Anand (**CSIR-CMERI**), Mukti Advani, Rina Singh (**CSIR-CRRI**), Suman Singh, Anamika Kothari (**CSIR-CSIO**), Avinash Mishra (**CSIR-CSMCRI**), Pooja Aggarwal (**CSIR-HQ**), Shreedhar Kanagarjan (**CSIR-HRDG**), Yogendra Padwad, Vikram Patial (**CSIR-IHBT**), Sumit G Gandhi, Fayaz Malik (**CSIR-IIIM**), Debashish Ghosh, Jyoti Porwal (**CSIR-IIP**), Vikas Srivastava, Prabhanshu Tripathi (**CSIR-IITR**), Anshu Bhardwaj, Srinivasan Krishnamurthi, Pradip Sen, Rashmi Kumar, Deepak Sharma, Amit Tuli, Pravin Kumar (**CSIR-IMTECH**), Indrani Ghosh (**IPU, CSIR Hqrs.**), Prakash L, Satisha Shri (**CSIR-NAL**), Chandana Venkateswara Rao, Sanjeev Kumar Ojha, Brahma Nanda Singh, Vijayanandraj Selvaraj (**CSIR-NBRI**), Shilpa Paranjape, Prashanti Niwant (**CSIR-NEERI**), Romi Wahengbam, Tridip Phukan, Pankaj Bharali (**CSIR-NEIST**), E V S S K Babu, Biswajit Mandal, T Vijaya Kumar (**CSIR-NGRI**), Rajeev K Sukumaran, Rameshkumar N (**CSIR-NIIST**), Samir Ravikant Damare, Kalpana Sandesh Chodankar, Bhumika Shirodkar (**CSIR-NIO**), Arvind Meena, Arun Uniyal (**CSIR-NIScPR**), Ansu J Kailath, Krishna Kumar, Roshan Kumar, Nikhil Kumar, Kuldeep Singh Gour (**CSIR-NML**), Ved Varun Agrawal, Arun Kant Singh (**CSIR-NPL**), Maheswaran Srinivasan, Vasudevan Pandurangan (**CSIR-SERC**), Rashmi Arya, Manisha Sakpal (**CSIR-URDIP**).

## Portal development and management

Kumardeep Chaudhary, Debasis Dash, Viren Sardana, Ajay Pratap Singh, Shantanu Sengupta, Satyartha Prakash, Vignesh S Kumar, Anshul Verma, Safeer Khan, Anshul Bhardwaj (**CSIR-IGIB**), Gopal Krishna Patra (**CSIR-4PI**), Nikhilesh Yadav (**CSIR-NCL**), Abbani Rakesh (**CSIR-NAL**).

## Sample Collection/Scanning

Pankaj Pandey, Vivek Junghare, Komal Jindal, Ankita Sahu, Satyartha Prakash, Meghana Arvind, Tarani Mathur, Shilpa Ray, Ayushi Narayan, Mamta Rathore, Yogesh Kumar, Vignesh S Kumar, Sreeshma Raj K, Anshul Verma, Pulkit Hasmukhbhai Leuva, Pratik Pathade, Safeer Khan, Anshul Bhardwaj, Shail Kumari, Bharti sharma, Ruchi, Shubham Kumar, Tanmay pawaskar, Sumant Kumar, Mohit, Shyam Singh Bisht, Deeksha Yadav, Rohit Kumar, Praveena Mishra, Ankit Basnal, Pranjal Tewari, Swati, Abhishek Kumar, Nancy Rawat, Mohammad Azhar Uddin, Deepak (**CSIR-IGIB**), MK Kanakavalli, Rakhesh K V, Neha Kumari, Dibya Rana Saha Roy (**CSIR-CCMB**), Ajit A Sutar, Sagar Baulia, Ameya A Pawar, Monika Sharma, Milind Kale, Ankita Namdeo, Rajesh S, Rashdajabeen Q Shaikh, Vaishnavi N Mahajan, Shivani V Palkar, Shrutika M Shewale, Shyam K Gawari (**CSIR-NCL**), Ankita Mridha, Saheli Chowdhury, Pratitusti Basu, Dipamoy Dutta, Dwaipayan Saha, (**CSIR-IICB**), Sai Adarsh Sahu, Sk Rameej Raja (**CSIR-IMMT**), Vishwa Priya (**CSIR-IICT**), Shail Singh, Lakra Promila, Rahul Roy,Shikha Yadav, Amit Kumar Shahravat, Kabita Sarkar, Adrija Rakshit, Deepanshu Sindhwani, Kajal KM, Smita Pandey (**CSIR-CDRI**), Vipul Sharma (**CSIR-CEERI**), Dikchha Singh (**CSIR-CIMFR**), Parimala Karupannan, Vandhana Anumaiya (**CSIR-CLRI**), Ravi Raj, Ankita Kumari (**CSIR-IHBT**), Nancy Sharma, Sahaurti Sharma, Sakshi Nagial, Kaneez Fatima (**CSIR-IIIM**), Suchismita Benjwal, Pramod Chauhan (**CSIR-IIP**), Neha Mehrotra (**CSIR-IITR**), Anshu Bhardwaj, Srinivasan Krishnamurthi, Parvez Ahmad, Priyadarshan Kinatukara, Bhupender Singh, Shiva Sundharam S, Pranavathiyani G, Kuldeep Singh, Rakesh Kumar, Siddhakam Palmal, Ritu Jatav, Lalit Kumar, Priyanshu Singh Raikwar, Simran Gambhir (**CSIR-IMTECH**), Madan Mohan Pandey (**CSIR-NBRI**), Manuj Kr Das, Borsha Rajkumari, Sukanya Borkakoti, Mamta Thapa, Ashique Hussain, Ishant Jyoti Nath (**CSIR-NEIST**), Akshika (**CSIR-NIScPR**), Navneet Singh Randhawa, Priyanka Singh, K Sudhakara Rao (**CSIR-NML**).

## Sample aliquoting

Aastha Mishra, Ankita Sahu, Meghana Arvind, Shivani Chitkara, Deeksha Yadav, Mohit, Shyam Singh Bisht, Shail Kumari, Ankit Basnal, Ansuman Sahu, Ruchi, Tanmay Pawaskar, Nancy Rawat, Pranjal Tewari, Vignesh S Kumar, Safeer Khan, Anshul Bhardwaj (**CSIR-IGIB**)

## Assays

Shantanu Sengupta, Viren Sardana, Kumardeep Chaudhary, Vamsi K. Yenamandra, Aastha Mishra, Ajay Pratap Singh, Ankita Sahu, Bharti Sharma, Shilpa Ray, Tarani Mathur, Mamta Rathore, Yogesh Kumar, Ankur Halder, Mohammad Azhar Uddin, Shubham Kumar, Sumant Kumar, Shail Kumari, Anshul Bhardwaj, Ruchi, Deeksha Yadav, Abhishek Kumar, Swati, Md. Intyaz Ali, Satyartha Prakash, Vignesh S Kumar, Safeer Khan (**CSIR-IGIB**), Partha Chakraborty, Dipyaman Ganguly, Saikat Majumder, Shilpak Chatterjee, Jahangir Alam (**CSIR-IICB**), Giriraj Ratan Chandak, Swasti Raychaudhuri, Karthik Bharadwaj Tallapaka, Jukanti Akshitha (**CSIR-CCMB**), Mahesh J Kulkarni, Chiranjit Chowdhury, Rajesh S, Ajit A Sutar, Ameya A Pawar, Milind Kale (**CSIR-NCL**), Umakanta Subudhi, Sai Adarsh Sahu, Sk Rameej Raja (**CSIR-IMMT**) Ramakrishna Sistla, Sai Balaji Andugulapati (**CSIR-IICT**), Amit Lahiri, Mrigank Srivastava, Shail Singh, Lakra Promila, Shikha Yadav, Kabita Sarkar, Adrija Rakshit, Deepanshu Sindhwani, Amit Kumar Shahravat (**CSIR-CDRI**)

## Data management and curation

Kumardeep Chaudhary, Viren Sardana, Shantanu Sengupta, Ajay Pratap Singh, Ankita Sahu, Meghana Arvind, Satyartha Prakash, Vignesh S Kumar, Pulkit Hasmukhbhai Leuva, Anshul Verma, Sreeshma Raj K, Pratik Pathade, Tarani Mathur, Kalyani Verma, Yogesh Kumar (**CSIR-IGIB**)

## Manuscript writing, review and editing

Viren Sardana, Shantanu Sengupta, Kumardeep Chaudhary, Anshul Verma, Meghana Arvind (**CSIR-IGIB**), Shalimar**\***, Sagnik Biswas**\* (AIIMS),** Partha Chakraborty (**CSIR-IICB**)

## Statistical Analysis and Data Visualization

Viren Sardana, Shantanu Sengupta, Kumardeep Chaudhary, Anshul Verma, Meghana Arvind, Sreeshma Raj K **(CSIR-IGIB),** Ashish Awasthi **(CDRI)**, Shalimar**\* (AIIMS),** Partha Chakrabarty **(IICB)**

## Phenome India consortium members not as authors

Rakesh Sharma, Beena Pillai, Vivek Rao, Sheetal Gandotra, Shantanu Chowdhury, Rajesh Pandey, Bhavana Prasher, Prateek Singh, Prajwali Sawant, Sudhir Rohilla, Rajat Ujjainiya, Harleen Kaur, Vishu Gupta, Mohit Kumar Divakar, Vikas M Hiremath, Saraswati Awasthi, Kanika Singh, Divya Bhalla, Anubhuti Bansal, Charvy Rana, Amit Maurya, Bharti, Shailja kant Upadhyay, Rushikesh Joshi, Shaivyanand Singh, Shivam Dhawan, Vanshika Srivastava, Dikkshita Baruah, Kamal Singh Pindari, Rohan Bhardwaj, Md, Quasid Akhter, Jitendra Kumar **(CSIR-IGIB),** Arun Bandyopadhyay, Ruby Banerjee **(CSIR-IICB),** Jatin Kalita, Prasenjit Manna, Rituraj Konwar, Himangsu Kousik, Bora Ravindra Kumar Rawal, Selvaraman Nagamani **(CSIR-NEIST),** Sumati Swain, Jukanti Akshitha, Deepshikha Esari, Mohammed Osed Annan, Mahfuj Hasan, Dimple Lavanuru **(CSIR-CCMB),** Srilekha Anumulapuri, Navya Sahithi Pelimelli, Abhisheik Eedara **(CSIR-IICT),** Swarnali Basu, Shweta Tiwari, Umesh Kumar (**CSIR-IIIM),** Arnold Moses, Rahul R Menon, Sanoj Mohan **(CSIR-NIIST)**

*Not consortium members

## Data Availability

Anonymized Data for public use may be made available after 3 years from completion of the baseline phase of the study or as per advisory from the Monitoring Committee of the project if any revisions are required.

## List of and Deidentified Abbreviations

MASLD: Metabolic Dysfunction Associated Steatotic Liver Disease
HbA1c: Glycated hemoglobin
RBC: Red Blood Cell
PCV: Packed cell volume
AST: Aspartate aminotransferase
ALT: Alanine aminotransferase
SGOT: Serum glutamic oxaloacetic transaminase
SGPT: Serum glutamic pyruvic transaminase
SLD: Steatotic liver disease
MAFLD: Metabolic Dysfunction-associated fatty liver disease
MASLD: Metabolic Dysfunction-associated fatty liver disease
CMRF: Cardiometabolic risk factor
MetALD: Metabolic and alcohol related/associated liver disease
NASH: Nonalcoholic steatohepatitis
HCC: Hepatocellular carcinoma
T2D: Type 2 Diabetes Mellites
LSM: Liver Stiffness Measurement
CAP: Controlled Attenuation Parameter
WC: Waist Circumference
HC: Hip Circumference
WHR: Waist Hip Ratio
HOMA-IR: Homeostatic Model of Insulin Resistance,
HDL: High-Density Lipoprotein
LDL: Low-Density Lipoprotein
E-Median: Liver Stiffness Measurement Score from Fibroscan
BMI: Body Mass Index

