## Supplementary Material for "Unveiling the Burden of MASLD and Liver Fibrosis in India: Novel Insights from the Phenome India Study into fibrosis without MASLD"

### Supplementary Section

### Methods

#### Anthropometry

As recommended by Integrated Child Development Services (ICDS) guidelines, a standard stadiometer was utilized at all centres. Weight (in kg) - Weight was measured using the Accuniq Body Composition Analyzer (Manufacturer- SELVAS Healthcare, Korea Model BC 380). Body Circumferences (in cm) - CESCORF measuring tape (Manufacturer – Cescorf, Brazil) was utilized to measure Chest circumference (CC), Waist circumference (WC), Abdominal circumference (AC), and Hip circumference (HC). The following definitions were adopted to measure the circumference three times at each site.

CC- “Measure the chest circumference at the most significant part of the chest, which is usually across the level of the nipple line in males and just above the breast tissue in females. Measurements were taken at the end of a normal expiration”.

WC- “A horizontal measure is taken at the midpoint between the lower margin of the last palpable rib and the top of the iliac crest”.

AC- “The tape is held behind the participant with one edge at the horizontal plane through the center of the umbilicus”.

HC- “The participant is standing erect, and the feet are close together. A horizontal measure is taken around the widest portion of the hips and buttocks.”

### Cytokine Analysis and Normalization of Data

Plasma samples were diluted at a ratio of 1:2 with the sample diluent supplied in the kit to optimize detection within the assay's dynamic range. The multiplex bead-based immunoassay was then conducted, which involved the sequential steps of bead incubation with diluted plasma, multiple wash cycles to remove unbound proteins, incubation with detection antibodies, and final incubation with streptavidin-phycoerythrin for signal generation. The assay plates were read using the Bio-Plex® 200 system (Bio-Rad Laboratories) and data were acquired with Bio-Plex Manager™ software. Standard curves were generated for each cytokine using the kit-provided standards and sample concentrations were calculated from these curves. A total of 4284 samples were analysed using 58 plates of the above-mentioned kit at two institutes, i.e., CSIR-IGIB (institute-1) and CSIR-IICB (institute-2). 33 plate runs were performed at institute-1, analysing 2391 samples, and 25 plate runs were performed at institute-2, analysing 893 samples. For normalization of data, analyte values corresponding to the control sample in each plate are extracted, and the median value is calculated for each analyte. The median value obtained in the previous step was used for computing the correction factor (CF) for each analyte in the respective plates. The equation below describes how the CF for an analyte on a particular plate is computed.

$$CF = \frac{Median\ value\ of\ analyte}{analyte\ value\ for\ control\ sample\ in\ particular\ plate}$$

Finally, the CF obtained for each analyte for respective plates is employed for scaling the analyte values in the corresponding plates using the equation below.

$$Normalized\ analyte\ value = original\ value * C$$

This method is commonly utilized in normalizing data for metabolite assays using PyComBat (21).

### Supplementary Figures

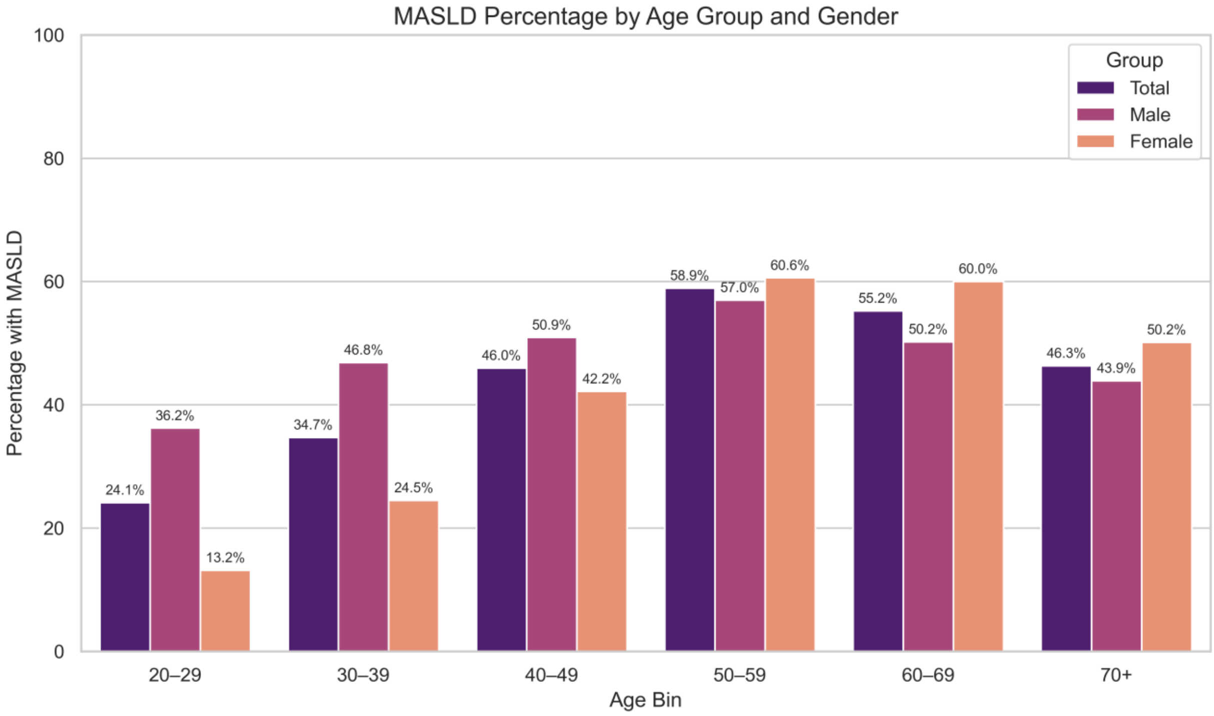


**Supplementary Figure 1:** Crude Age and Gender based distribution of MASLD in Phenome India Data

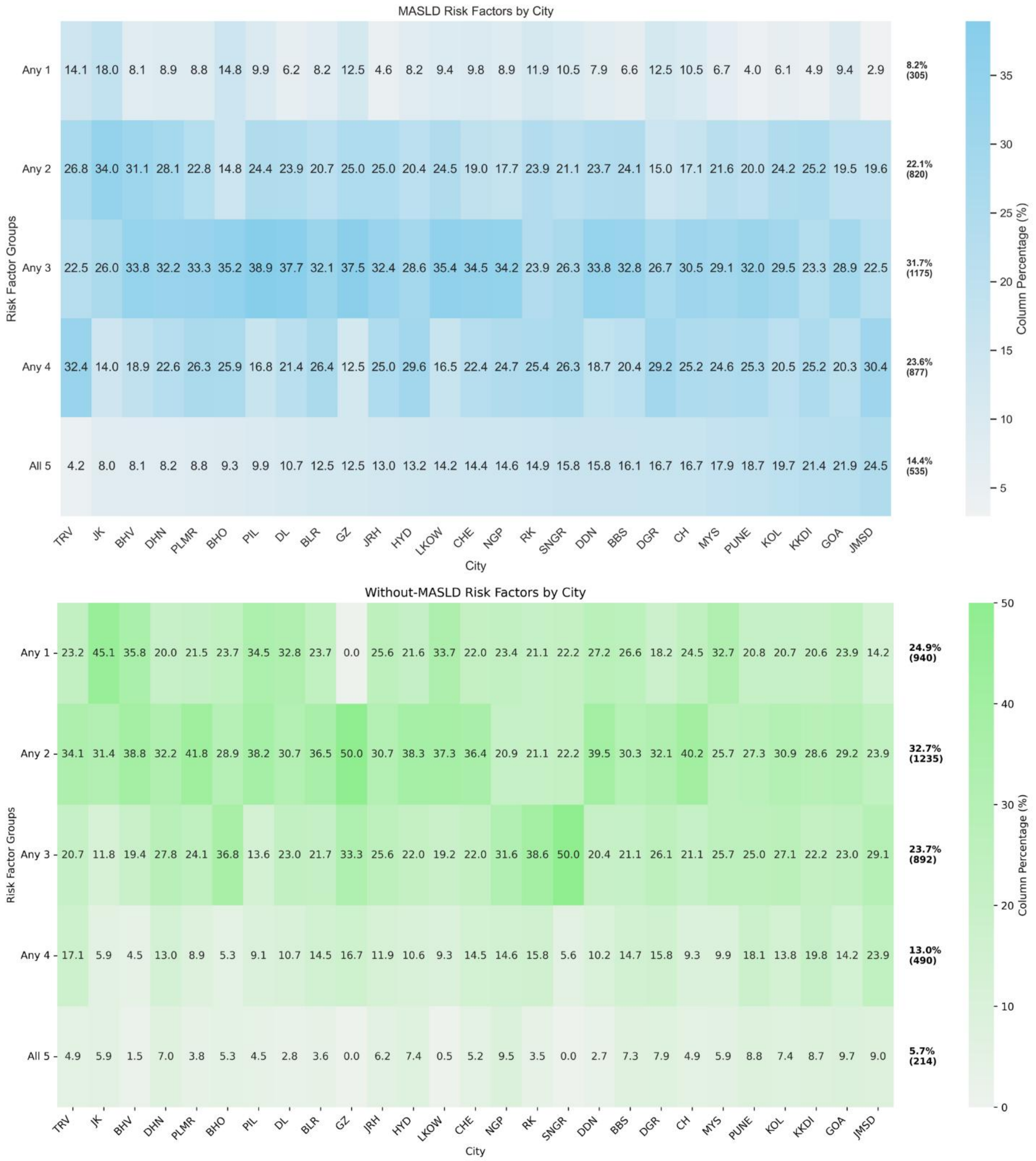


**Supplementary Figure 2:** Prevalence of CMRF in MASLD population by city within MASLD and without-MASLD groups. Data are presented as percentages within age groups of CMRFs (vertical). Outside figure percentages and numbers reflect the total MASLD and the without-MASLD group contribution. Without-MASLD, it may not add to the complete number for other participants who had zero risk factors (n=281).

Abbreviations: TRV, Thiruvananthapuram; KOL, Kolkata; DGR, Durgapur; DDN, Dehradun; PLMR, Palampur; KKDI, Karaikudi; CH, Chandigarh; PUNE, Pune; JMSD, Jamshedpur; BLR, Bengaluru; JRH, Jorhat; DL, Delhi; LKOW, Lucknow; MYS, Mysore; NGP, Nagpur; PIL, Pilani; RK, Roorkee; JK, Jammu; GOA, Goa; DHN, Dhanbad; BBS, Bhubaneswar; HYD, Hyderabad; CHE, Chennai; BHV, Bhavnagar; BHO, Bhopal; GZ, Ghaziabad; SNGR, Srinagar.

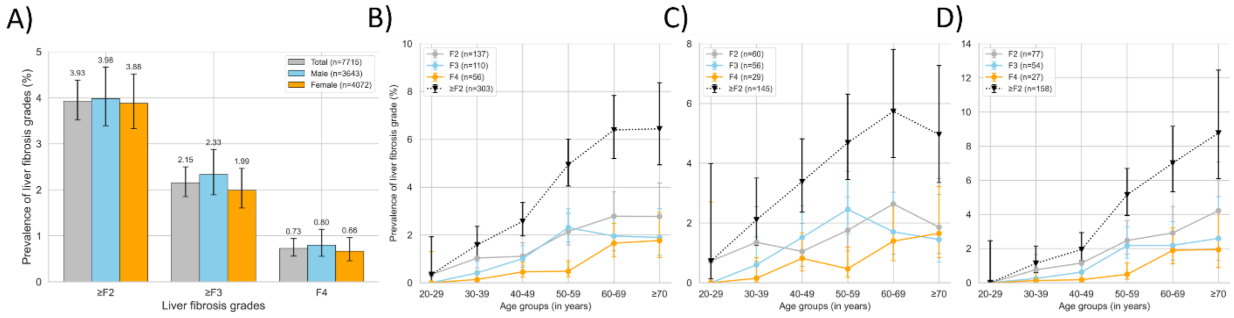


**Supplementary Figure 3:** Crude Prevalence of liver fibrosis grades for all participants (n=7715), gender and age (B-D, Total, male and female respectively) (Data is presented as prevalence (proportions) with 95% confidence intervals (error bars). Data are presented as prevalence proportions with 95% confidence intervals.

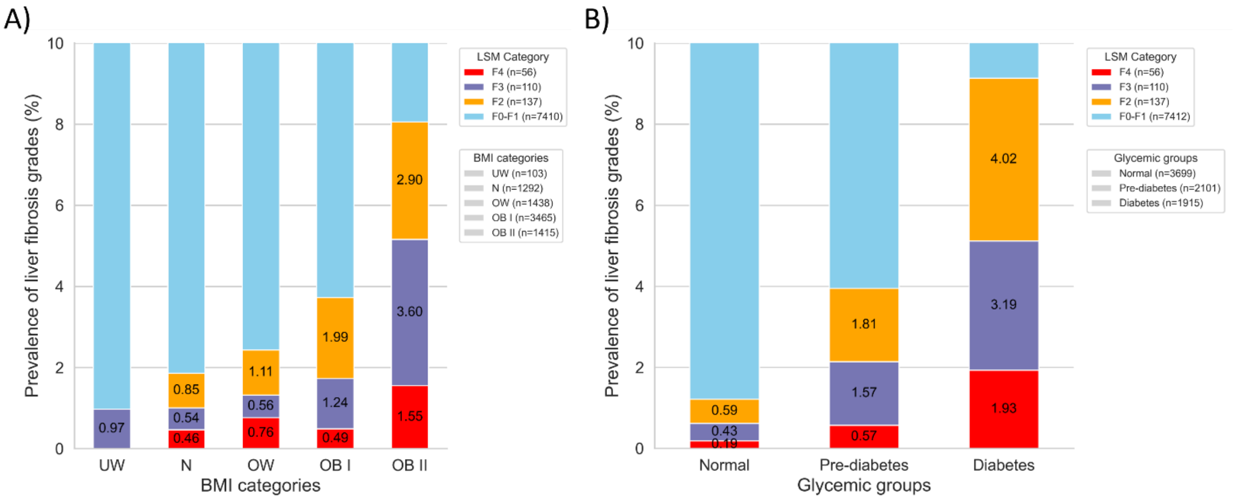


**Supplementary Figure 4:** Prevalence of liver fibrosis as per different BMI sub-groups (n=7713) and in participants with diabetes and pre-diabetics (n=7715). A) Data based on BMI for the distribution of F0-F1, F2, F3 and F4. B) Data based on Glycaemia control (Normal, Prediabetic, Diabetic) for the distribution of F0-F1, F2, F3 and F4.

Abbreviations: UW-Underweight; N- Normal/Healthy weight; OW-Overweight; OB I-Obese I, OB II-Obese II.

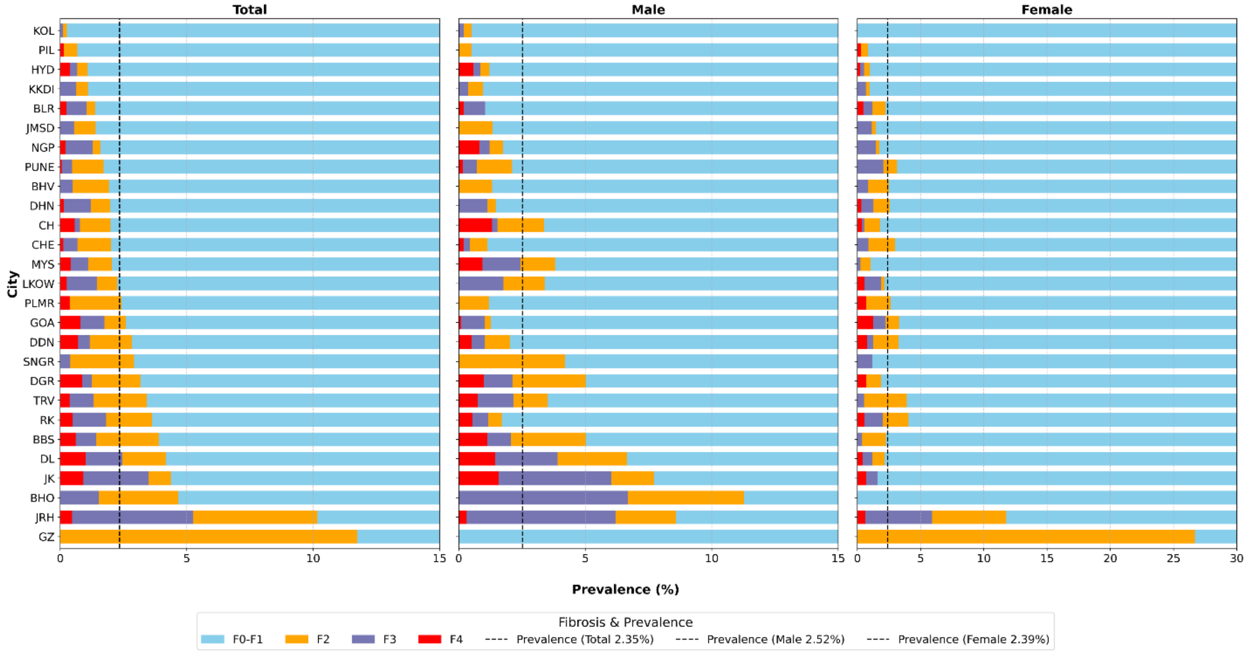


**Supplementary Figure 5:** City/District-wise age-adjusted prevalence of F2, F3, F4 fibrosis in Phenome India Cohort. The Dotted Line indicates the age-adjusted prevalence for the overall group, with the value in parentheses (Data in Supplementary Table 5).

Abbreviations: TRV, Thiruvananthapuram; KOL, Kolkata; DGR, Durgapur; DDN, Dehradun; PLMR, Palampur; KKDI, Karaikudi; CH, Chandigarh; PUNE, Pune; JMSD, Jamshedpur; BLR, Bengaluru; JRH, Jorhat; DL, Delhi; LKOW, Lucknow; MYS, Mysore; NGP, Nagpur; PIL, Pilani; RK, Roorkee; JK, Jammu; GOA, Goa; DHN, Dhanbad; BBS, Bhubaneswar; HYD, Hyderabad; CHE, Chennai; BHV, Bhavnagar; BHO, Bhopal; GZ, Ghaziabad; SNGR, Srinagar.

**Supplementary Table 1:** Demographics of Phenome India Cohort

| Variable | n / Median (% / IQR) | | |
|---|---|---|---|
| | Total | Male | Female |
| Sample size | 10267 | 5840 (56.88) | 4427 (43.12) |
| Age (years) | 50.0 (41.0-61.0) | 51.0 (41.0-62.0) | 49.0 (40.0-59.0) |
| Aspartate Aminotransferase (U/L) | 24.60 (20.60-30.50) | 26.30 (22.0-32.60) | 22.60 (19.10-27.60) |
| Alanine Aminotransferase (U/L) | 22.90 (16.70-33.20) | 26.90 (19.60-38.50) | 18.90 (14.40-26.05) |
| Glucose Fasting (mg/dL) | 91.30 (84.60-103.50) | 92.50 (85.30-105.80) | 89.7 (83.60-100.80) |
| Glycosylated Haemoglobin (%) | 5.60 (5.30-6.20) | 5.70 (5.30-6.30) | 5.60 (5.30-6.10) |
| Diabetes | 2648 (25.80) | 1644 (28.15) | 1003 (22.66) |
| Homa IR | 1.89 (1.29-2.79) | 1.95 (1.32-2.88) | 1.83 (1.26-2.66) |
| Total Cholesterol (mg/dL) | 175.95 (150.70-202.40) | 175.0 (149.30-201.20) | 177.45 (153.08-204.20) |
| Serum HDL cholesterol (mg/dL) | 47.80 (41.60-54.70) | 45.30 (39.90-51.50) | 51.25 (44.90-58.80) |
| Serum LDL cholesterol (mg/dL) | 114.90 (95.0-135.50) | 115.30 (94.70-136.30) | 114.40 (95.40-134.60) |
| Serum Triglycerides (mg/dL) | 113.90 (85.60-154.08) | 120.30 (90.80-163.33) | 105.95 (79.50-140.70) |
| Elevated Blood Pressure | 5812 (56.60) | 3515 (60.19) | 2297 (51.89) |
| Haemoglobin (g/dL) | 13.40 (12.20-14.50) | 14.30 (13.40-15.10) | 12.30 (11.40-13.0) |
| **Anthropometry Parameters** | | | |
| Weight (kg) | 68.50 (61.28-76.58) | 71.40 (64.50-79.70) | 64.32 (57.8-71.9) |
| Height (cm) | 162.0 (155.0-168.4) | 167.40 (162.93-171.90) | 154.2 (150.2-158.4) |
| Waist circumference (cm) | 88.23 (81.83-95.33) | 89.43 (84.07-95.90) | 85.83 (78.67-94.33) |
| Hip circumference (cm) | 97.83 (92.93-103.70) | 96.17 (92.0-101.0) | 101.05 (94.97-107.49) |
| Body Mass Index (kg/m$^2$) | 26.10 (23.80-28.90) | 25.50 (23.50-27.98) | 27.10 (24.40-30.10) |
| Waist-Hip Ratio | 0.90 (0.84-0.96) | 0.93 (0.89-0.98) | 0.85 (0.79-0.90) |
| **BMI (n=10261)** | | | |
| Lean | 1792 (17.46) | 1153 (19.76) | 639 (14.43) |
| Overweight | 1950 (19.00) | 1312 (22.48) | 638 (14.41) |
| Obese I | 4670 (45.51) | 2671 (45.77) | 1999 (45.15) |
| Obese II | 1849 (18.02) | 700 (11.99) | 1149 (25.95) |

Continuous variables are summarized using median (Q1, Q3), and categorical variables are summarized using n (%); missing data for each variable were excluded when producing these summaries. Lean includes underweight: BMI <18.5 kg/m² and normal/healthy weight: BMI 18.5–22.9 kg/m².

Abbreviations: Homa IR, Homeostatic Model Assessment of Insulin Resistance; HDL, High-density lipoprotein; LDL, Low-density lipoprotein; CAP, Controlled Attenuation Parameter.

**Supplementary Table 2:** Comparison of relevant history, diet and socio-economic status between MASLD and without-MASLD population subgroups

| **Variable Name** | **MASLD n (%)** | **Without-MASLD n (%)** | **Adjusted P-Value (FDR)** |
|---|---|---|---|
| **Female with menopause** | 1119 (30.1%) | 810 (20.0%) | 1.442e-49 |
| **Liver disease** | 263 (7.1%) | 150 (3.7%) | 5.933e-11 |
| **Physical activity** | 2045 (55.1%) | 2149 (53.0%) | 0.082 |
| **Educational Qualification** | | | |
| *Doctorate* | 857 (23.1%) | 903 (22.3%) | |
| *Master's degree* | 1011 (27.2%) | 1204 (29.7%) | |
| *Bachelor's degree* | 961 (25.9%) | 1097 (27.1%) | |
| *Secondary education or high school* | 575 (15.5%) | 607 (15.0%) | |
| *Primary education* | 221 (6.0%) | 174 (4.3%) | |
| *No formal education* | 87 (2.3%) | 66 (1.6%) | |
| **Monthly Income** | | | |
| *Greater than 300,000* | 583 (15.7%) | 588 (14.5%) | |
| *200,000 - 300,000* | 379 (10.2%) | 352 (8.7%) | |
| *100,000 - 200,000* | 899 (24.2%) | 1026 (25.3%) | |
| *50,000 - 100,000* | 903 (24.3%) | 1000 (24.7%) | |
| *25,000 - 50,000* | 568 (15.3%) | 616 (15.2%) | |
| *Less than 25,000* | 380 (10.2%) | 469 (11.6%) | |
| **Diet type** | | | |
| *Non-vegetarian* | 1869 (50.4%) | 2128 (52.5%) | |
| *Vegetarian (dairy)* | 1211 (32.6%) | 1228 (30.3%) | |
| *Vegetarian (dairy + egg)* | 402 (10.8%) | 464 (11.5%) | |
| *Vegan (no dairy or egg products)* | 142 (3.8%) | 136 (3.4%) | |
| *Vegetarian (egg)* | 88 (2.4%) | 96 (2.4%) | |

Continuous variables are summarized using n (%); missing data for each variable were excluded when producing these summaries.

**Supplementary Table 3**: Multivariate models to assess the contribution of factors influencing and contributing to MASLD

| | OR | Robust SE | z | P>\|z\| | 95% LCI | 95% UCI |
|---|---|---|---|---|---|---|
| **Age Group (Against 20-30)** | | | | | | |
| 30-40 | 1.10 | 0.14 | 0.79 | 0.43 | 0.86 | 1.41 |
| 40-50 | 1.33 | 0.18 | 2.10 | 0.04 | 1.02 | 1.73 |
| 50-60 | 1.72 | 0.25 | 3.76 | 0.00 | 1.30 | 2.29 |
| 60-70 | 1.33 | 0.19 | 1.99 | 0.05 | 1.00 | 1.75 |
| 70+ | 1.09 | 0.16 | 0.59 | 0.55 | 0.82 | 1.45 |
| **Gender** | | | | | | |
| Female | 0.60 | 0.04 | -6.86 | 0.00 | 0.52 | 0.70 |
| **Diabetes Status** | | | | | | |
| Yes | 2.24 | 0.16 | 11.61 | 0.00 | 1.96 | 2.57 |
| **Hypertension Status** | | | | | | |
| Yes | 1.29 | 0.08 | 3.94 | 0.00 | 1.14 | 1.46 |
| **Dyslipidemia** | | | | | | |
| Yes | 1.65 | 0.08 | 10.37 | 0.00 | 1.50 | 1.82 |
| **BMI Group (Against Normal)** | | | | | | |
| Underweight | 0.20 | 0.12 | -2.75 | 0.01 | 0.06 | 0.63 |
| Overweight | 2.63 | 0.23 | 10.98 | 0.00 | 2.22 | 3.13 |
| Obese_I | 5.62 | 0.57 | 16.95 | 0.00 | 4.60 | 6.86 |
| Obese_II | 13.77 | 1.30 | 27.76 | 0.00 | 11.44 | 16.57 |
| **Waist Hip Ratio** | | | | | | |
| Above .80/.90 | 1.61 | 0.11 | 6.79 | 0.00 | 1.40 | 1.84 |
| Constant | 0.08 | 0.01 | -16.48 | 0.00 | 0.06 | 0.11 |

**Supplementary Table 4**: Comparison of MASLD subtypes

| Variable Name | **Lean MASLD**<br>**Median (IQR)**<br>***n (%)*** | **Overweight MASLD**<br>**Median (IQR)**<br>***n (%)*** | **Obese MASLD**<br>**Median (IQR)**<br>***n (%)*** | **Adjusted P-Value (FDR)** |
|---|---|---|---|---|
| **n (%)** | 233 (6.3%) | 530 (14.3%) | 2949 (79.4%) | |
| **Sex (Male)** | 149 (63.9%) | 367 (69.2%) | 1319 (44.7%) | |
| **Sex (Female)** | 84 (36.1%) | 163 (30.8%) | 1630 (55.3%) | |
| **Age (years)** | 58.00 (49.00–67.00)<br>*233 (6.3%)* | 53.00 (44.00–62.00)<br>*530 (14.3%)* | 52.00 (44.00–61.00)<br>*2949 (79.4%)* | 1.206e-07 |
| **Aspartate Aminotransferase (AST/SGOT)** | 25.40 (21.20–29.30)<br>*233 (6.3%)* | 26.20 (21.50–33.00)<br>*529 (14.3%)* | 25.00 (20.70–31.70)<br>*2940 (79.4%)* | 3.858e-02 |
| **Alanine Aminotransferase (ALT/SGPT)** | 24.30 (17.90–32.70)<br>*233 (6.3%)* | 27.10 (19.20–38.80)<br>*529 (14.3%)* | 24.80 (18.10–36.45)<br>*2940 (79.4%)* | 1.476e-02 |
| **Glucose, Fasting** | 98.80 (88.40–121.70)<br>*233 (6.3%)* | 93.60 (86.00–113.35)<br>*527 (14.2%)* | 94.80 (86.80–110.35)<br>*2939 (79.5%)* | 6.757e-03 |
| **Total Cholesterol** | 184.00 (156.30–212.70)<br>*233 (6.3%)* | 176.30 (152.70–206.50)<br>*529 (14.3%)* | 179.70 (154.00–205.95)<br>*2944 (79.4%)* | 0.181 |
| **Serum HDL Cholesterol** | 49.00 (43.30–56.20)<br>*233 (6.3%)* | 47.00 (40.50–53.30)<br>*529 (14.3%)* | 47.50 (41.40–54.00)<br>*2944 (79.4%)* | 3.372e-02 |
| **Serum LDL Cholesterol** | 120.60 (97.80–142.90)<br>*233 (6.3%)* | 118.50 (96.60–139.60)<br>*529 (14.3%)* | 118.25 (98.10–139.05)<br>*2944 (79.4%)* | 0.455 |
| **Serum Triglycerides** | 131.70 (95.80–168.40)<br>*233 (6.3%)* | 129.30 (95.70–174.40)<br>*529 (14.3%)* | 126.30 (98.05–167.15)<br>*2944 (79.4%)* | 0.670 |
| **Hba1c (Glycosylated Hemoglobin)** | 6.00 (5.50–7.25)<br>*230 (6.3%)* | 5.80 (5.40–6.70)<br>*525 (14.3%)* | 5.90 (5.50–6.60)<br>*2919 (79.5%)* | 3.312e-02 |
| **Haemoglobin** | 13.60 (12.50–14.70)<br>*231 (6.3%)* | 13.85 (12.80–14.70)<br>*522 (14.2%)* | 13.10 (12.00–14.20)<br>*2921 (79.5%)* | 7.840e-19 |
| **HOMA IR** | 1.82 (1.22–2.48)<br>*228 (6.3%)* | 2.07 (1.48–2.94)<br>*508 (14.1%)* | 2.47 (1.75–3.55)<br>*2855 (79.5%)* | 1.482e-21 |
| **Dyslipidaemia** | 151 (64.8%) | 305 (57.5%) | 1676 (56.8%) | 0.073 |

| | | | | |
|---|---|---|---|---|
| **Diabetes** | 99 (42.5%) | 179 (33.8%) | 1002 (34.0%) | 3.858e-02 |
| **Elevated blood pressure** | 156 (67.0%) | 305 (57.5%) | 1925 (65.3%) | 4.791e-03 |
| **Female with menopause** | 65 (27.9%) | 103 (19.4%) | 951 (32.2%) | 4.791e-03 |

Continuous variables are summarized using median (Q1, Q3), and categorical variables are summarized using n (%); missing data for each variable were excluded when producing these summaries.

Abbreviations: HDL, High-density lipoprotein; LDL, Low-density lipoprotein; Homa IR, Homeostatic Model Assessment of Insulin Resistance.

**Supplementary Table 5:** Age-adjusted Prevalence of MASLD by City

| City | Total | Male | Female |
|---|---|---|---|
| **TRV (162)** | 25.34 (20.67 - 30.01) | 31.15 (22.90 - 39.40) | 25.61 (19.76 - 31.46) |
| **KOL (334)** | 29.88 (24.75 - 35.00) | 31.88 (24.48 - 39.28) | 33.07 (24.46 - 41.67) |
| **DDN (293)** | 31.63 (26.26 - 36.99) | 43.96 (30.06 - 57.85) | 28.99 (24.22 - 33.76) |
| **DGR (299)** | 32.11 (26.63 - 37.59) | 39.88 (29.87 - 49.90) | 30.73 (23.67 - 37.79) |
| **PLMR (145)** | 33.20 (26.77 - 39.63) | 60.84 (52.03 - 69.64) | 31.24 (24.60 - 37.87) |
| **KKDI (234)** | 33.85 (27.69 - 40.00) | 34.20 (25.13 - 43.28) | 31.71 (23.61 - 39.82) |
| **NGP (329)** | 34.21 (28.67 - 39.76) | 52.09 (39.55 - 64.64) | 32.49 (27.22 - 37.75) |
| **BLR (641)** | 35.37 (31.37 - 39.36) | 41.63 (35.78 - 47.48) | 30.81 (25.97 - 35.65) |
| **CH (425)** | 35.74 (31.04 - 40.43) | 44.68 (36.63 - 52.73) | 34.96 (28.21 - 41.70) |
| **PUNE (458)** | 35.74 (30.72 - 40.76) | 51.03 (39.10 - 62.96) | 32.83 (27.74 - 37.93) |
| **JRH (292)** | 35.97 (28.68 - 43.25) | 42.69 (30.29 - 55.10) | 31.70 (25.35 - 38.04) |
| **JMSD (243)** | 36.36 (27.15 - 45.57) | 50.35 (41.65 - 59.06) | 33.04 (26.12 - 39.95) |
| **DL (708)** | 36.92 (33.15 - 40.70) | 53.76 (47.30 - 60.22) | 28.36 (24.30 - 32.42) |
| **LKOW (419)** | 38.36 (32.36 - 44.37) | 57.71 (46.77 - 68.64) | 29.63 (21.98 - 37.28) |
| **JK (104)** | 38.97 (30.39 - 47.56) | 68.40 (54.30 - 82.50) | 19.60 (10.78 - 28.42) |
| **PIL (266)** | 39.17 (31.90 - 46.44) | 41.03 (31.08 - 50.99) | 33.70 (27.72 - 39.68) |
| **MYS (239)** | 39.95 (33.02 - 46.88) | 58.91 (45.24 - 72.58) | 35.35 (27.10 - 43.59) |
| **GOA (251)** | 41.11 (34.48 - 47.73) | 43.55 (33.92 - 53.19) | 35.89 (27.31 - 44.47) |
| **DHN (271)** | 41.45 (35.22 - 47.67) | 50.40 (39.60 - 61.21) | 38.23 (29.09 - 47.36) |
| **BBS (251)** | 42.03 (36.86 - 47.20) | 46.03 (38.05 - 54.00) | 36.45 (30.19 - 42.72) |
| **CHE (358)** | 42.76 (36.54 - 48.99) | 52.00 (43.00 - 61.00) | 33.36 (25.62 - 41.10) |
| **HYD (612)** | 42.88 (35.09 - 50.68) | 49.23 (38.70 - 59.76) | 34.14 (29.10 - 39.19) |
| **BHV (155)** | 46.92 (37.23 - 56.62) | 47.09 (36.35 - 57.84) | 55.75 (46.73 - 64.77) |
| **RK (130)** | 48.19 (33.25 - 63.13) | 63.79 (51.39 - 76.20) | 32.95 (18.03 - 47.86) |
| **BHO (94)** | 49.38 (36.90 - 61.86) | 71.65 (58.87 - 84.44) | 36.61 (22.32 - 50.89) |
| **GZ (14)** | 52.87 (24.45 - 81.29) | 18.06 (9.28 - 26.84) | 88.36 (72.24 - 100.00) |
| **SNGR (37)** | 57.43 (36.33 - 78.53) | 64.20 (45.17 - 83.22) | 59.49 (34.55 - 84.44) |

Values shown as Prevalence (95% CI)

Abbreviations: TRV, Thiruvananthapuram; KOL, Kolkata; DGR, Durgapur; DDN, Dehradun; PLMR, Palampur; KKDI, Karaikudi; CH, Chandigarh; PUNE, Pune; JMSD, Jamshedpur; BLR, Bengaluru; JRH, Jorhat; DL, Delhi; LKOW, Lucknow; MYS, Mysore; NGP, Nagpur; PIL, Pilani; RK, Roorkee; JK, Jammu; GOA, Goa; DHN, Dhanbad; BBS, Bhubaneswar; HYD, Hyderabad; CHE, Chennai; BHV, Bhavnagar; BHO, Bhopal; GZ, Ghaziabad; SNGR, Srinagar.

**Supplementary Table 6:** Age-adjusted Prevalence of Fibrosis by City

| City | F0-F1 | F2 | F3 | F4 |
|---|---|---|---|---|
| **KOL (333)** | 99.74 (99.37 - 100.00) | 0.16 (0.00 - 0.46) | 0.11 (0.00 - 0.31) | 0.00 (0.00 - 0.00) |
| **PIL (266)** | 99.32 (98.67 - 99.98) | 0.53 (0.00 - 1.14) | 0.00 (0.00 - 0.00) | 0.14 (0.00 - 0.42) |
| **HYD (611)** | 98.91 (98.27 - 99.56) | 0.41 (0.01 - 0.81) | 0.28 (0.00 - 0.61) | 0.39 (0.00 - 0.78) |
| **KKDI (234)** | 98.91 (97.75 - 100.00) | 0.46 (0.00 - 1.18) | 0.64 (0.00 - 1.54) | 0.00 (0.00 - 0.00) |
| **BLR (641)** | 98.62 (97.87 - 99.37) | 0.33 (0.00 - 0.69) | 0.80 (0.23 - 1.36) | 0.25 (0.00 - 0.59) |
| **JMSD (240)** | 98.60 (97.41 - 99.79) | 0.85 (0.00 - 1.80) | 0.55 (0.00 - 1.31) | 0.00 (0.00 - 0.00) |
| **NGP (328)** | 98.42 (96.99 - 99.84) | 0.29 (0.00 - 0.70) | 1.08 (0.00 - 2.39) | 0.21 (0.00 - 0.60) |
| **PUNE (454)** | 98.29 (97.23 - 99.34) | 1.25 (0.27 - 2.22) | 0.41 (0.00 - 0.83) | 0.06 (0.00 - 0.18) |
| **BHV (153)** | 98.08 (95.96 - 100.00) | 1.44 (0.00 - 3.33) | 0.49 (0.00 - 1.42) | 0.00 (0.00 - 0.00) |
| **DHN (271)** | 98.02 (96.89 - 99.15) | 0.78 (0.04 - 1.51) | 1.05 (0.21 - 1.90) | 0.15 (0.00 - 0.43) |
| **CH (425)** | 98.01 (96.87 - 99.15) | 1.21 (0.27 - 2.14) | 0.21 (0.00 - 0.50) | 0.57 (0.00 - 1.18) |
| **CHE (357)** | 97.99 (96.67 - 99.31) | 1.33 (0.23 - 2.43) | 0.56 (0.00 - 1.26) | 0.12 (0.00 - 0.35) |
| **MYS (238)** | 97.96 (96.53 - 99.40) | 0.94 (0.00 - 1.90) | 0.69 (0.00 - 1.44) | 0.41 (0.00 - 1.20) |
| **LKOW (417)** | 97.75 (96.66 - 98.84) | 0.80 (0.17 - 1.43) | 1.19 (0.35 - 2.02) | 0.26 (0.00 - 0.62) |
| **PLMR (145)** | 97.58 (94.28 - 100.00) | 2.04 (0.00 - 5.27) | 0.00 (0.00 - 0.00) | 0.37 (0.00 - 1.10) |
| **GOA (251)** | 97.40 (95.98 - 98.82) | 0.86 (0.00 - 1.84) | 0.96 (0.08 - 1.83) | 0.79 (0.18 - 1.41) |
| **DDN (293)** | 97.18 (95.69 - 98.66) | 1.66 (0.51 - 2.80) | 0.46 (0.00 - 0.97) | 0.71 (0.00 - 1.53) |
| **SNGR (37)** | 97.08 (92.74 - 100.00) | 2.52 (0.00 - 6.80) | 0.40 (0.00 - 1.13) | 0.00 (0.00 - 0.00) |
| **DGR (289)** | 96.83 (95.20 - 98.46) | 1.92 (0.58 - 3.27) | 0.37 (0.00 - 0.88) | 0.87 (0.05 - 1.70) |
| **TRV (162)** | 96.59 (94.82 - 98.36) | 2.09 (0.75 - 3.42) | 0.96 (0.00 - 2.02) | 0.37 (0.00 - 1.07) |
| **RK (130)** | 96.36 (93.39 - 99.34) | 1.82 (0.00 - 4.35) | 1.33 (0.00 - 2.76) | 0.49 (0.00 - 1.13) |
| **BBS (242)** | 96.11 (94.10 - 98.11) | 2.48 (0.86 - 4.10) | 0.80 (0.00 - 1.68) | 0.61 (0.00 - 1.47) |
| **DL (702)** | 95.81 (94.21 - 97.41) | 1.74 (0.47 - 3.00) | 1.45 (0.69 - 2.21) | 1.00 (0.36 - 1.65) |
| **JK (104)** | 95.62 (92.53 - 98.72) | 0.88 (0.00 - 2.52) | 2.59 (0.11 - 5.06) | 0.92 (0.00 - 2.15) |
| **BHO (92)** | 95.33 (91.88 - 98.77) | 3.14 (1.10 - 5.19) | 1.53 (0.00 - 4.30) | 0.00 (0.00 - 0.00) |
| **JRH (286)** | 89.83 (86.30 - 93.36) | 4.91 (2.16 - 7.66) | 4.79 (2.44 - 7.14) | 0.47 (0.05 - 0.89) |
| **GZ (14)** | 88.26 (72.00 - 100.00) | 11.74 (0.00 - 28.00) | 0.00 (0.00 - 0.00) | 0.00 (0.00 - 0.00) |

Values shown as Prevalence (95% CI) for each LSM category (F0-F4)

Abbreviations: TRV, Thiruvananthapuram; KOL, Kolkata; DGR, Durgapur; DDN, Dehradun; PLMR, Palampur; KKDI, Karaikudi; CH, Chandigarh; PUNE, Pune; JMSD, Jamshedpur; BLR, Bengaluru; JRH, Jorhat; DL, Delhi; LKOW, Lucknow; MYS, Mysore; NGP, Nagpur; PIL, Pilani; RK, Roorkee; JK, Jammu; GOA, Goa; DHN, Dhanbad; BBS, Bhubaneswar; HYD, Hyderabad; CHE, Chennai; BHV, Bhavnagar; BHO, Bhopal; GZ, Ghaziabad; SNGR, Srinagar.

**Supplementary Table 7:** Multivariate models to assess the contribution of factors influencing and contributing to fibrosis

| | OR | Robust SE | z | P>\|z\| | 95% LCI | 95% UCI |
|---|---|---|---|---|---|---|
| **Age Group (Against 20-30)** | | | | | | |
| 30-40 | 2.173257 | 1.194685 | 1.41 | 0.158 | 0.739925 | 6.383139 |
| 40-50 | 3.015176 | 2.141492 | 1.55 | 0.12 | 0.749475 | 12.13021 |
| 50-60 | 3.929731 | 2.804429 | 1.92 | 0.055 | 0.970304 | 15.91541 |
| 60-70 | 4.777081 | 3.574192 | 2.09 | 0.037 | 1.102293 | 20.70275 |
| 70+ | 4.981876 | 3.53398 | 2.26 | 0.024 | 1.240447 | 20.00818 |
| **Gender** | | | | | | |
| Female | 0.804341 | 0.158263 | -1.11 | 0.268 | 0.546961 | 1.182834 |
| **Diabetes Status** | | | | | | |
| Yes | 3.12287 | 0.433636 | 8.2 | 0 | 2.378798 | 4.099683 |
| **Hypertension Status** | | | | | | |
| Yes | 1.085893 | 0.133336 | 0.67 | 0.502 | 0.853628 | 1.381354 |
| **Dyslipedemia** | | | | | | |
| Yes | 0.78963 | 0.108655 | -1.72 | 0.086 | 0.602971 | 1.034071 |
| **BMI Group (Against Normal)** | | | | | | |
| Obese_I | 1.836157 | 0.592198 | 1.88 | 0.06 | 0.975843 | 3.454932 |
| Obese_II | 4.271986 | 1.754107 | 3.54 | 0 | 1.910384 | 9.552984 |
| Over_weight | 1.2321 | 0.380585 | 0.68 | 0.499 | 0.672538 | 2.257225 |
| Under_weight | 0.736025 | 0.844196 | -0.27 | 0.789 | 0.077731 | 6.969346 |
| **Waist-Hip Ratio** | | | | | | |
| Above .80/.90 | 2.173493 | 0.558728 | 3.02 | 0.003 | 1.313242 | 3.59726 |
| Constant | 0.002342 | 0.001536 | -9.23 | 0 | 0.000648 | 0.008471 |

**Supplementary Table 8:** Age-adjusted Prevalence of MASLD by Zone

| Zone | Total | Male | Female |
|---|---|---|---|
| **West (1193)** | 38.64 (34.93 - 42.34) | 43.59 (36.72 - 50.46) | 36.29 (32.15 - 40.44) |
| **North (1685)** | 37.66 (34.38 - 40.94) | 45.45 (39.61 - 51.30) | 29.97 (27.15 - 32.78) |
| **South (2246)** | 37.46 (33.89 - 41.03) | 44.88 (38.07 - 51.69) | 31.42 (28.78 - 34.06) |
| **Central (950)** | 36.60 (32.84 - 40.35) | 54.52 (46.38 - 62.65) | 29.82 (25.54 - 34.09) |
| **North-East (292)** | 35.97 (28.68 - 43.25) | 42.69 (30.29 - 55.10) | 31.70 (25.35 - 38.04) |
| **East (1398)** | 34.35 (31.69 - 37.01) | 37.23 (33.32 - 41.15) | 31.55 (28.06 - 35.04) |

Values shown as Prevalence (95% CI)

**Supplementary Table 9:** Age-adjusted Prevalence of Fibrosis by Zone

| Zone | F0-F1 | F2 | F3 | F4 |
|---|---|---|---|---|
| **South (2243)** | 98.53 (98.15 - 98.92) | 0.64 (0.39 - 0.88) | 0.60 (0.34 - 0.86) | 0.23 (0.08 - 0.39) |
| **East (1375)** | 98.10 (97.55 - 98.64) | 1.04 (0.63 - 1.45) | 0.58 (0.27 - 0.88) | 0.28 (0.07 - 0.49) |
| **West (1186)** | 97.71 (96.82 - 98.60) | 1.25 (0.49 - 2.01) | 0.72 (0.30 - 1.14) | 0.32 (0.10 - 0.54) |
| **Central (946)** | 97.28 (96.47 - 98.08) | 1.36 (0.77 - 1.94) | 0.96 (0.47 - 1.44) | 0.41 (0.11 - 0.71) |
| **North (1679)** | 97.12 (96.34 - 97.91) | 1.36 (0.73 - 1.99) | 0.91 (0.52 - 1.29) | 0.61 (0.33 - 0.89) |
| **North-East (286)** | 89.83 (86.30 - 93.36) | 4.91 (2.16 - 7.66) | 4.79 (2.44 - 7.14) | 0.47 (0.05 - 0.89) |

Values shown as Prevalence (95% CI) for each LSM category (F0-F4)

**Supplementary Table 10**: Comparison of Anthropometric differences between MASLD and without MASLD associated fibrosis population subgroups

| Variable Name | MASLD [With fibrosis] Median (IQR) *n (%)* | Without-MASLD [With fibrosis] Median (IQR) *n (%)* | Adjusted P-Value (FDR) Median (IQR) *n (%)* |
|---|---|---|---|
| **Weight (kg)** | 74.70 (66.15–86.50)<br>*234 (77.2%)* | 65.10 (57.60–70.80)<br>*69 (22.8%)* | 7.659e-09 |
| **Height (cm)** | 159.90 (151.90–166.55)<br>*234 (77.2%)* | 157.10 (152.00–166.00)<br>*69 (22.8%)* | 0.421 |
| **Body Mass Index (kg/m$^2$)** | 29.60 (26.60–33.40)<br>*234 (77.2%)* | 25.10 (23.00–28.40)<br>*69 (22.8%)* | 5.770e-10 |
| **Waist-Hip ratio** | 0.94 (0.88–0.99)<br>*229 (77.1%)* | 0.91 (0.86–0.96)<br>*68 (22.9%)* | 1.208e-02 |
| **Body Mass Index Category** | | | 2.684e-11 |
| **Underweight (<18.5)** | *0 (0.0%)* | *1 (1.4%)* | |
| **Normal (18.5-22.9)** | *8 (3.4%)* | *16 (23.2%)* | |
| **Overweight (23-24.9)** | *18 (7.7%)* | *17 (24.6%)* | |
| **Obese I** | *102 (43.6%)* | *27 (39.1%)* | |
| **Obese II** | *106 (45.3%)* | *8 (11.6%)* | |
| **FibroScan® LSM (in kPa)** | | | 0.138 |
| **F0-F1** | | | |
| **F2** | *106 (45.3%)* | *31 (44.9%)* | |
| **F3** | *90 (38.5%)* | *20 (29.0%)* | |
| **F4** | *38 (16.2%)* | *18 (26.1%)* | |
| **CAP Enhanced Mean** | 307.50 (285.00–330.00)<br>*234 (77.2%)* | 216.00 (198.00–232.00)<br>*69 (22.8%)* | 2.060e-35 |

Continuous variables are summarized using median (Q1, Q3) and categorical variables are summarized using n (%); missing data for each variable were excluded when producing these summaries.

Abbreviations: LSM, Liver stiffness measurement; CAP, Controlled Attenuation parameter

**Supplementary Table 11:** Comparison of cytokine levels across MASLD and *w*ithout-MASLD population subgroups

| Variable Name | MASLD<br>Median (IQR) [n] | Without MASLD<br>Median (IQR) [n] | Adjusted P-Value (FDR) |
|---|---|---|---|
| **b-NGF** | 0.53 (0.24–0.96) [1130] | 0.47 (0.22–0.90) [1051] | 0.210 |
| **CTACK** | 153.96 (97.92–232.11) [1626] | 151.79 (86.58–230.50) [1613] | 0.173 |
| **Eotaxin** | 19.01 (10.53–29.99) [1647] | 16.72 (8.33–28.10) [1655] | 3.120e-04 |
| **FGF basic** | 35.44 (23.89–50.18) [1414] | 33.52 (23.26–47.60) [1375] | 4.721e-02 |
| **G-CSF** | 26.30 (14.35–41.33) [1421] | 22.55 (12.76–35.35) [1350] | 1.437e-06 |
| **GM-CSF** | 2.11 (0.97–2.68) [99] | 1.70 (0.88–2.47) [108] | 0.173 |
| **GRO-a** | 428.81 (292.75–585.57) [1463] | 419.73 (285.65–565.18) [1438] | 0.260 |
| **HGF** | 178.07 (127.17–238.66) [1646] | 159.23 (111.70–209.28) [1647] | 1.850e-12 |
| **IFN-a2** | 2.70 (1.63–4.74) [806] | 2.91 (1.68–5.04) [752] | 0.421 |
| **IFN-g** | 11.82 (8.06–17.78) [1606] | 10.70 (7.46–15.80) [1588] | 8.918e-05 |
| **IL-1a** | 14.42 (8.80–22.15) [1508] | 13.35 (6.78–21.05) [1468] | 2.102e-03 |
| **IL-1b** | 5.44 (3.51–8.03) [1633] | 4.64 (3.01–6.81) [1611] | 5.596e-12 |
| **IL-1ra** | 107.94 (65.67–171.69) [1475] | 94.93 (61.39–153.63) [1420] | 2.519e-04 |
| **IL-2** | 1.44 (0.69–2.32) [828] | 1.37 (0.69–2.31) [802] | 0.626 |
| **IL-2Ra** | 30.83 (20.96–41.73) [1645] | 28.41 (19.54–38.44) [1653] | 4.926e-04 |
| **IL-3** | 0.10 (0.05–0.21) [836] | 0.10 (0.04–0.18) [829] | 0.247 |
| **IL-4** | 0.86 (0.56–1.18) [1483] | 0.82 (0.51–1.14) [1472] | 1.545e-02 |
| **IL-5** | 337.16 (67.42–390.73) [41] | 319.63 (38.36–410.69) [52] | 0.642 |
| **IL-6** | 1.10 (0.61–1.78) [1145] | 0.97 (0.50–1.73) [1069] | 8.560e-03 |
| **IL-7** | 18.44 (11.37–29.21) [1467] | 16.14 (10.14–25.42) [1453] | 4.219e-06 |
| **IL-8** | 2.36 (1.47–3.78) [1568] | 2.11 (1.29–3.32) [1559] | 1.437e-06 |
| **IL-9** | 226.50 (132.48–316.33) [1634] | 214.38 (120.28–311.73) [1642] | 3.990e-02 |
| **IL-10** | 5.24 (2.91–8.29) [1321] | 5.24 (2.80–8.26) [1280] | 0.421 |
| **IL-12(p70)** | 1.19 (0.56–2.13) [715] | 1.07 (0.51–2.10) [716] | 0.586 |
| **IL-12(p40)** | 17.43 (8.54–30.98) [1055] | 17.24 (8.23–31.13) [1015] | 0.911 |
| **IL-13** | 1.39 (0.79–2.55) [1465] | 1.32 (0.75–2.55) [1423] | 0.301 |

| | | | |
|---|---|---|---|
| **IL-15** | 126.56 (74.66–190.56) [173] | 139.95 (91.07–190.56) [160] | 0.287 |
| **IL-16** | 30.11 (21.59–40.77) [1618] | 28.74 (19.73–38.43) [1612] | 1.110e-03 |
| **IL-17A** | 3.21 (2.07–4.78) [1443] | 3.02 (1.87–4.56) [1436] | 3.868e-02 |
| **IL-18** | 29.41 (18.62–41.82) [1648] | 25.54 (16.11–37.70) [1646] | 2.548e-07 |
| **IP-10** | 84.29 (45.61–147.58) [1613] | 71.31 (36.42–127.30) [1613] | 1.764e-06 |
| **LIF** | 34.31 (21.01–48.36) [1451] | 31.58 (19.84–48.55) [1383] | 0.155 |
| **M-CSF** | 6.92 (4.31–9.97) [1616] | 6.14 (3.59–9.21) [1596] | 1.437e-06 |
| **MCP-1(MCAF)** | 7.09 (4.39–10.43) [1609] | 6.22 (3.87–9.56) [1599] | 1.574e-05 |
| **MCP-3** | 2.03 (1.12–3.32) [1047] | 1.97 (1.13–3.12) [960] | 0.439 |
| **MIF** | 333.05 (213.70–482.52) [1637] | 317.91 (194.28–484.93) [1635] | 0.064 |
| **MIG** | 154.73 (100.73–259.98) [1606] | 145.73 (88.21–269.38) [1604] | 0.063 |
| **MIP-1a** | 0.72 (0.50–1.03) [1624] | 0.63 (0.44–0.87) [1621] | 1.566e-12 |
| **MIP-1b** | 87.92 (63.04–113.87) [1649] | 84.62 (59.94–109.78) [1654] | 1.540e-02 |
| **PDGF-bb** | 33.40 (16.80–61.34) [1488] | 31.03 (16.00–59.15) [1485] | 0.335 |
| **RANTES** | 1359.31 (601.77–2500.55) [1654] | 1267.47 (523.06–2449.47) [1664] | 3.282e-02 |
| **SCF** | 40.82 (28.03–55.32) [1613] | 37.87 (25.66–54.64) [1613] | 8.848e-03 |
| **SCGF-b** | 24840.47 (15533.54–35592.13) [1646] | 23898.84 (14771.37–34536.86) [1649] | 0.055 |
| **SDF-1a** | 387.13 (235.28–555.85) [1598] | 399.61 (235.49–580.06) [1593] | 0.260 |
| **TNF-a** | 28.61 (17.49–41.38) [1604] | 27.10 (16.12–41.04) [1595] | 0.063 |
| **TNF-b** | 143.14 (89.07–197.78) [1646] | 135.89 (80.17–197.78) [1649] | 0.064 |
| **TRAIL** | 19.21 (11.97–26.12) [1624] | 17.73 (11.08–24.04) [1611] | 2.381e-04 |
| **VEGF** | 42.71 (30.63–81.11) [88] | 40.01 (27.53–87.09) [98] | 0.725 |

Continuous variables are summarized using median (Q1, Q3), and categorical variables are summarized using n (%); missing data for each variable were excluded when producing these summaries.

**Supplementary Table 12**: Cytokine levels in different fibrosis subgroups with and without MASLD

| Cytokines | MASLD without fibrosis Median (IQR) *n (%)* | Without-MASLD without fibrosis Median (IQR) *n (%)* | Adjusted P-Value (FDR) | MASLD with fibrosis Median (IQR) *n (%)* | Without-MASLD with fibrosis Median (IQR) *n (%)* | Adjusted P-Value (FDR) |
|---|---|---|---|---|---|---|
| **N (%)** | 1447 (47.5%) | 1599 (52.5%) | | 203 (76.3%) | 63 (23.7%) | |
| **b-NGF** | 0.47 (0.21 - 0.81) [952] | 0.46 (0.20 - 0.85) [993] | 0.891 | 1.03 (0.57 - 1.66) [170] | 1.89 (1.14 - 2.41) [52] | 6.23E-04 |
| **CTACK** | 147.74 (91.59 - 215.92) [1413] | 147.80 (84.41 - 218.82) [1537] | 0.85 | 239.76 (142.23 - 372.66) [202] | 288.77 (200.81 - 387.44) [63] | 0.154 |
| **Eotaxin** | 18.30 (9.49 - 28.68) [1434] | 16.30 (8.10 - 27.14) [1579] | 4.33E-03 | 25.90 (16.05 - 41.62) [202] | 43.02 (27.88 - 66.29) [63] | 3.14E-04 |
| **FGF basic** | 34.01 (22.81 - 45.46) [1222] | 32.97 (22.73 - 46.13) [1304] | 0.779 | 54.65 (37.96 - 67.51) [182] | 52.60 (38.57 - 67.22) [59] | 0.777 |
| **G-CSF** | 24.76 (13.67 - 38.92) [1217] | 21.87 (12.31 - 33.60) [1277] | 6.70E-05 | 39.06 (23.77 - 59.35) [193] | 59.04 (42.17 - 80.90) [61] | 3.14E-04 |
| **GM-CSF** | 2.19 (0.80 - 3.03) [77] | 2.04 (0.98 - 2.50) [90] | 0.463 | 2.00 (1.10 - 2.61) [21] | 0.92 (0.42 - 1.52) [18] | 0.05 |
| **GRO-a** | 416.35 (286.67 - 565.93) [1267] | 412.66 (284.23 - 558.77) [1372] | 0.779 | 526.96 (371.01 - 694.82) [187] | 580.79 (448.00 - 781.32) [56] | 0.15 |
| **HGF** | 172.15 (121.52 - 225.92) [1433] | 156.40 (109.82 - 205.36) [1571] | 9.23E-08 | 240.57 (175.83 - 340.93) [202] | 280.50 (205.73 - 409.71) [63] | 0.123 |
| **IFN-a2** | 2.50 (1.60 - 4.65) [682] | 2.86 (1.66 - 4.82) [703] | 0.374 | 3.58 (1.98 - 5.43) [119] | 4.95 (2.88 - 7.38) [43] | 0.153 |
| **IFN-g** | 11.23 (7.72 - 15.93) [1397] | 10.58 (7.33 - 15.28) [1512] | 0.065 | 27.89 (12.34 - 36.26) [198] | 31.70 (13.80 - 41.49) [63] | 0.312 |
| **IL-1a** | 14.22 (8.21 - 22.00) [1306] | 12.69 (6.52 - 20.40) [1395] | 3.91E-03 | 16.01 (10.75 - 22.50) [193] | 25.17 (16.01 - 33.15) [61] | 8.24E-04 |
| **IL-1b** | 5.25 (3.37 - 7.69) [1421] | 4.59 (2.91 - 6.77) [1536] | 6.06E-07 | 7.87 (4.92 - 10.69) [201] | 5.66 (4.47 - 7.39) [63] | 1.26E-03 |
| **IL-1ra** | 104.82 (64.57 - 163.08) [1275] | 92.67 (59.47 - 151.30) [1351] | 7.61E-03 | 150.24 (98.77 - 205.12) [191] | 159.90 (108.04 - 199.61) [58] | 0.528 |
| **IL-2** | 1.29 (0.66 - 2.12) [676] | 1.26 (0.66 - 2.12) [748] | 0.918 | 2.16 (1.15 - 3.50) [146] | 4.98 (3.50 - 6.83) [47] | 5.93E-06 |
| **IL-2Ra** | 29.54 (20.28 - 40.23) [1432] | 28.13 (19.26 - 37.82) [1577] | 2.28E-02 | 37.18 (27.14 - 49.55) [202] | 37.68 (28.41 - 50.00) [63] | 0.528 |
| **IL-3** | 0.08 (0.04 - 0.16) [708] | 0.08 (0.04 - 0.16) [794] | 0.779 | 0.22 (0.09 - 0.39) [150] | 0.42 (0.20 - 0.63) [52] | 1.42E-03 |
| **IL-4** | 0.84 (0.55 - 1.18) [1285] | 0.80 (0.50 - 1.13) [1401] | 0.092 | 0.98 (0.77 - 1.31) [187] | 1.08 (0.77 - 1.71) [60] | 0.131 |
| **IL-5** | 353.57 (255.07 - 397.62) [29] | 319.63 (112.80 - 417.22) [50] | 0.494 | 257.22 (59.93 - 323.77) [11] | 23.03 (11.98 - 34.08) [2] | 0.154 |
| **IL-6** | 1.03 (0.56 - 1.75) [965] | 0.95 (0.49 - 1.70) [1009] | 0.143 | 1.34 (0.87 - 1.92) [172] | 1.23 (0.83 - 2.10) [50] | 0.694 |
| **IL-7** | 17.93 (11.20 - 26.44) [1271] | 15.82 (9.96 - 25.28) [1379] | 3.91E-03 | 33.48 (16.27 - 45.00) [186] | 26.88 (18.30 - 34.73) [61] | 0.154 |
| **IL-8** | 2.21 (1.39 - 3.35) [1358] | 2.04 (1.25 - 3.15) [1484] | 3.91E-03 | 4.50 (2.67 - 6.22) [199] | 5.45 (2.89 - 7.87) [62] | 4.54E-02 |
| **IL-9** | 221.33 (129.54 - 316.80) [1423] | 212.74 (120.61 - 310.25) [1569] | 0.195 | 247.05 (175.23 - 314.67) [202] | 247.50 (135.84 - 331.79) [60] | 0.836 |
| **IL-10** | 5.30 (2.78 - 8.29) [1137] | 5.12 (2.77 - 8.26) [1208] | 0.445 | 5.18 (3.07 - 8.24) [177] | 6.39 (4.56 - 9.13) [60] | 0.129 |

| | | | | | | |
|---|---|---|---|---|---|---|
| **IL-12(p70)** | 1.09 (0.52 - 2.02) [589] | 1.01 (0.47 - 1.99) [674] | 0.374 | 1.68 (0.60 - 2.81) [122] | 3.22 (2.17 - 4.60) [44] | 3.14E-04 |
| **IL-12(p40)** | 16.27 (7.67 - 29.64) [882] | 16.87 (7.94 - 30.87) [956] | 0.494 | 29.26 (14.80 - 43.07) [163] | 34.09 (22.15 - 60.59) [50] | 4.83E-02 |
| **IL-13** | 1.37 (0.76 - 2.44) [1265] | 1.30 (0.71 - 2.43) [1353] | 0.374 | 1.57 (0.93 - 3.32) [190] | 2.86 (1.79 - 4.58) [58] | 2.17E-03 |
| **IL-15** | 135.73 (99.00 - 200.79) [105] | 132.66 (87.73 - 190.56) [128] | 0.494 | 88.86 (40.28 - 157.67) [66] | 157.67 (126.56 - 189.37) [31] | 2.22E-03 |
| **IL-16** | 29.22 (21.12 - 39.83) [1409] | 28.30 (19.51 - 37.50) [1537] | 2.70E-02 | 35.93 (26.96 - 50.18) [198] | 41.26 (32.61 - 49.63) [62] | 0.15 |
| **IL-17A** | 3.14 (2.00 - 4.61) [1241] | 2.98 (1.87 - 4.48) [1363] | 0.154 | 3.97 (2.44 - 5.54) [192] | 5.68 (3.20 - 7.49) [61] | 2.26E-02 |
| **IL-18** | 28.36 (17.70 - 40.42) [1435] | 25.05 (15.90 - 36.40) [1570] | 2.29E-05 | 34.99 (26.79 - 49.16) [202] | 41.39 (31.88 - 54.07) [63] | 0.129 |
| **IP-10** | 79.32 (42.51 - 135.37) [1403] | 69.06 (35.84 - 122.59) [1538] | 7.39E-04 | 146.00 (78.80 - 256.73) [199] | 244.49 (115.86 - 351.66) [63] | 2.48E-02 |
| **LIF** | 33.42 (20.52 - 48.70) [1254] | 31.22 (19.36 - 48.65) [1315] | 0.189 | 34.46 (25.11 - 43.33) [188] | 38.59 (29.35 - 45.78) [58] | 0.119 |
| **M-CSF** | 6.73 (4.21 - 9.69) [1404] | 6.06 (3.54 - 9.07) [1524] | 1.02E-04 | 8.81 (6.19 - 11.87) [201] | 9.18 (6.34 - 13.17) [62] | 0.489 |
| **MCP-1(MCAF)** | 6.72 (4.17 - 9.81) [1398] | 6.05 (3.81 - 9.22) [1525] | 5.33E-03 | 10.39 (7.26 - 14.15) [200] | 11.02 (8.43 - 15.32) [62] | 0.242 |
| **MCP-3** | 2.03 (1.19 - 3.35) [914] | 1.98 (1.13 - 3.13) [916] | 0.208 | 1.48 (0.89 - 3.00) [129] | 1.90 (1.37 - 2.94) [37] | 0.216 |
| **MIF** | 324.91 (205.56 - 473.34) [1425] | 318.31 (191.16 - 485.57) [1560] | 0.374 | 375.01 (269.95 - 530.67) [201] | 311.54 (235.81 - 438.62) [62] | 0.103 |
| **MIG** | 146.26 (97.77 - 254.10) [1396] | 141.83 (86.52 - 252.55) [1530] | 0.224 | 204.38 (143.10 - 357.83) [200] | 323.38 (185.23 - 601.40) [62] | 2.17E-03 |
| **MIP-1a** | 0.68 (0.47 - 0.98) [1412] | 0.62 (0.43 - 0.85) [1547] | 9.23E-08 | 1.05 (0.78 - 1.42) [201] | 1.21 (0.95 - 1.67) [61] | 0.103 |
| **MIP-1b** | 86.42 (61.47 - 111.45) [1436] | 84.03 (60.14 - 109.09) [1579] | 0.2 | 101.46 (73.87 - 126.63) [202] | 95.38 (49.71 - 132.46) [63] | 0.383 |
| **PDGF-bb** | 33.10 (17.40 - 59.42) [1295] | 30.79 (15.86 - 59.07) [1410] | 0.374 | 33.82 (14.59 - 70.64) [183] | 38.01 (17.00 - 78.61) [62] | 0.512 |
| **RANTES** | 1337.90 (585.53 - 2454.73) [1440] | 1279.04 (542.06 - 2448.76) [1588] | 0.2 | 1530.48 (695.12 - 2996.34) [203] | 1117.75 (307.37 - 2339.69) [63] | 4.54E-02 |
| **SCF** | 39.65 (27.11 - 53.56) [1403] | 37.63 (25.44 - 53.34) [1537] | 0.143 | 51.72 (35.64 - 65.83) [199] | 56.76 (37.50 - 68.02) [63] | 0.299 |
| **SCGF-b** | 23670.32 (14501.39 - 34718.04) [1433] | 23530.16 (14418.37 - 34022.08) [1574] | 0.374 | 31410.78 (23658.04 - 41067.07) [202] | 32974.34 (24850.41 - 41733.84) [63] | 0.844 |
| **SDF-1a** | 386.24 (231.62 - 549.63) [1389] | 394.10 (230.18 - 565.29) [1517] | 0.494 | 389.59 (254.55 - 594.52) [198] | 811.33 (429.59 - 1141.08) [63] | 5.55E-06 |
| **TNF-a** | 27.33 (16.98 - 40.37) [1395] | 26.95 (15.96 - 40.66) [1520] | 0.297 | 36.70 (24.17 - 46.89) [198] | 40.64 (26.54 - 52.64) [62] | 0.249 |
| **TNF-b** | 144.38 (87.71 - 201.05) [1434] | 136.40 (81.50 - 199.00) [1574] | 0.198 | 138.88 (105.07 - 182.06) [201] | 131.69 (72.68 - 174.39) [63] | 0.117 |
| **TRAIL** | 19.02 (11.47 - 25.88) [1414] | 17.72 (10.88 - 24.24) [1537] | 7.67E-03 | 21.64 (16.22 - 27.16) [200] | 17.80 (14.79 - 22.38) [62] | 2.48E-02 |
| **VEGF** | 42.36 (32.19 - 83.94) [59] | 40.01 (30.79 - 83.49) [74] | 0.779 | 47.32 (24.82 - 72.69) [29] | 42.18 (24.82 - 85.33) [22] | 0.871 |

Continuous variables are summarized using median (Q1, Q3), and categorical variables are summarized using n (%); missing data for each variable were excluded when producing these summaries.

**Supplementary Table 13:** Comparison of cytokine levels across MASLD with and without fibrosis.

| Cytokines | MASLD with fibrosis | MASLD without fibrosis | Adjusted P-Value (FDR) |
|---|---|---|---|
| **N (%)** | 203 (12.3%) | 1447 (87.7%) | |
| **b-NGF** | 1.03 (0.57 - 1.66) [170] | 0.47 (0.21 - 0.81) [952] | **4.92E-20** |
| **CTACK** | 239.76 (142.23 - 372.66) [202] | 147.74 (91.59 - 215.92) [1413] | **4.12E-18** |
| **Eotaxin** | 25.90 (16.05 - 41.62) [202] | 18.30 (9.49 - 28.68) [1434] | **1.14E-10** |
| **FGF basic** | 54.65 (37.96 - 67.51) [182] | 34.01 (22.81 - 45.46) [1222] | **1.94E-20** |
| **G-CSF** | 39.06 (23.77 - 59.35) [193] | 24.76 (13.67 - 38.92) [1217] | **3.42E-13** |
| **GM-CSF** | 2.00 (1.10 - 2.61) [21] | 2.19 (0.80 - 3.03) [77] | 0.8913 |
| **GRO-a** | 526.96 (371.01 - 694.82) [187] | 416.35 (286.67 - 565.93) [1267] | **5.03E-08** |
| **HGF** | 240.57 (175.83 - 340.93) [202] | 172.15 (121.52 - 225.92) [1433] | **2.81E-22** |
| **IFN-a2** | 3.58 (1.98 - 5.43) [119] | 2.50 (1.60 - 4.65) [682] | **0.0199** |
| **IFN-g** | 27.89 (12.34 - 36.26) [198] | 11.23 (7.72 - 15.93) [1397] | **9.85E-31** |
| **IL-1a** | 16.01 (10.75 - 22.50) [193] | 14.22 (8.21 - 22.00) [1306] | **0.0071** |
| **IL-1b** | 7.87 (4.92 - 10.69) [201] | 5.25 (3.37 - 7.69) [1421] | **3.86E-13** |
| **IL-1ra** | 150.24 (98.77 - 205.12) [191] | 104.82 (64.57 - 163.08) [1275] | **1.00E-08** |
| **IL-2** | 2.16 (1.15 - 3.50) [146] | 1.29 (0.66 - 2.12) [676] | **1.28E-08** |
| **IL-2Ra** | 37.18 (27.14 - 49.55) [202] | 29.54 (20.28 - 40.23) [1432] | **2.07E-09** |
| **IL-3** | 0.22 (0.09 - 0.39) [150] | 0.08 (0.04 - 0.16) [708] | **1.56E-19** |
| **IL-4** | 0.98 (0.77 - 1.31) [187] | 0.84 (0.55 - 1.18) [1285] | **2.05E-05** |
| **IL-5** | 257.22 (59.93 - 323.77) [11] | 353.57 (255.07 - 397.62) [29] | 0.0808 |
| **IL-6** | 1.34 (0.87 - 1.92) [172] | 1.03 (0.56 - 1.75) [965] | **2.59E-05** |
| **IL-7** | 33.48 (16.27 - 45.00) [186] | 17.93 (11.20 - 26.44) [1271] | **6.33E-17** |
| **IL-8** | 4.50 (2.67 - 6.22) [199] | 2.21 (1.39 - 3.35) [1358] | **1.27E-29** |
| **IL-9** | 247.05 (175.23 - 314.67) [202] | 221.33 (129.54 - 316.80) [1423] | 0.0584 |
| **IL-10** | 5.18 (3.07 - 8.24) [177] | 5.30 (2.78 - 8.29) [1137] | 0.6618 |
| **IL-12(p70)** | 1.68 (0.60 - 2.81) [122] | 1.09 (0.52 - 2.02) [589] | **0.0047** |
| **IL-12(p40)** | 29.26 (14.80 - 43.07) [163] | 16.27 (7.67 - 29.64) [882] | **7.98E-10** |
| **IL-13** | 1.57 (0.93 - 3.32) [190] | 1.37 (0.76 - 2.44) [1265] | **0.0059** |
| **IL-15** | 88.86 (40.28 - 157.67) [66] | 135.73 (99.00 - 200.79) [105] | **0.0035** |
| **IL-16** | 35.93 (26.96 - 50.18) [198] | 29.22 (21.12 - 39.83) [1409] | **1.75E-09** |
| **IL-17A** | 3.97 (2.44 - 5.54) [192] | 3.14 (2.00 - 4.61) [1241] | **5.04E-06** |
| **IL-18** | 34.99 (26.79 - 49.16) [202] | 28.36 (17.70 - 40.42) [1435] | **2.83E-08** |
| **IP-10** | 146.00 (78.80 - 256.73) [199] | 79.32 (42.51 - 135.37) [1403] | **4.12E-18** |
| **LIF** | 34.46 (25.11 - 43.33) [188] | 33.42 (20.52 - 48.70) [1254] | 0.7303 |
| **M-CSF** | 8.81 (6.19 - 11.87) [201] | 6.73 (4.21 - 9.69) [1404] | **1.59E-08** |
| **MCP-1(MCAF)** | 10.39 (7.26 - 14.15) [200] | 6.72 (4.17 - 9.81) [1398] | **4.72E-19** |
| **MCP-3** | 1.48 (0.89 - 3.00) [129] | 2.03 (1.19 - 3.35) [914] | **0.0087** |
| **MIF** | 375.01 (269.95 - 530.67) [201] | 324.91 (205.56 - 473.34) [1425] | **0.001** |

| | | | |
|---|---|---|---|
| **MIG** | 204.38 (143.10 - 357.83) [200] | 146.26 (97.77 - 254.10) [1396] | **9.72E-09** |
| **MIP-1a** | 1.05 (0.78 - 1.42) [201] | 0.68 (0.47 - 0.98) [1412] | **8.54E-25** |
| **MIP-1b** | 101.46 (73.87 - 126.63) [202] | 86.42 (61.47 - 111.45) [1436] | **2.94E-05** |
| **PDGF-bb** | 33.82 (14.59 - 70.64) [183] | 33.10 (17.40 - 59.42) [1295] | 0.8489 |
| **RANTES** | 1530.48 (695.12 - 2996.34) [203] | 1337.90 (585.53 - 2454.73) [1440] | 0.0736 |
| **SCF** | 51.72 (35.64 - 65.83) [199] | 39.65 (27.11 - 53.56) [1403] | **7.98E-10** |
| **SCGF-b** | 31410.78 (23658.04 - 41067.07) [202] | 23670.32 (14501.39 - 34718.04) [1433] | **1.14E-09** |
| **SDF-1a** | 389.59 (254.55 - 594.52) [198] | 386.24 (231.62 - 549.63) [1389] | 0.0966 |
| **TNF-a** | 36.70 (24.17 - 46.89) [198] | 27.33 (16.98 - 40.37) [1395] | **4.09E-06** |
| **TNF-b** | 138.88 (105.07 - 182.06) [201] | 144.38 (87.71 - 201.05) [1434] | 0.9835 |
| **TRAIL** | 21.64 (16.22 - 27.16) [200] | 19.02 (11.47 - 25.88) [1414] | **0.0011** |
| **VEGF** | 47.32 (24.82 - 72.69) [29] | 42.36 (32.19 - 83.94) [59] | 0.5362 |

Continuous variables are summarized using median (Q1, Q3), and categorical variables are summarized using n (%); missing data for each variable were excluded when producing these summaries.
